# Animals in higher education settings: Do animal-assisted interventions improve mental and cognitive health outcomes of students? A systematic review and meta-analysis

**DOI:** 10.1101/2022.04.11.22273607

**Authors:** Annalena Huber, Stefanie J. Klug, Annette Abraham, Erica Westenberg, Veronika Schmidt, Andrea S. Winkler

## Abstract

**Background:** Due to the high burden of mental health issues among students at higher education institutions world-wide, animal-assisted interventions (AAIs) are being increasingly used to relieve student stress. The objective of this study was to systematically review of the effects of AAIs on the mental and cognitive health outcomes of higher education students.

**Methods:** Randomized controlled trials using any unfamiliar animal as the sole intervention tool were included in the systematic review. Study quality was assessed using the Cochrane Risk-of-Bias tool. Where possible, effect sizes (Hedges’ g) were pooled for individual outcomes using random-effects meta-analyses. Albatross plots were used to supplement the data synthesis.

**Results:** Of 2.401 identified studies, 35 were included. Almost all studies used dogs as the intervention animal. The quality of most included studies was rated as moderate. Studies showed an overall reduction of acute anxiety (g= -0.57 (95%CI -1.45;0.31)) and stress. For other mental outcomes, studies showed an overall small reduction of negative affect (g= -0.47 (95%CI -1.46;0.52)), chronic stress (g= -0.23 (95%CI -0.57;0.11)) and depression, as well as small increases in arousal, happiness and positive affect (g= 0.06 (95%CI -0.78;0.90)). Studies showed no effect on heart rate and heart rate variability, a small reduction in salivary cortisol and mixed effects on blood pressure. No effect on cognitive outcomes was found.

**Conclusion:** Overall, evidence suggests that AAIs are effective at improving mental, but not physiological or cognitive outcomes of students. Strong methodological heterogeneity between studies limited the ability to draw clear conclusions.

## Introduction

As highlighted by ongoing events such as climate change and the COVID-19 pandemic, which is strongly suspected to have zoonotic origins [1], it is essential to acknowledge the interconnectedness of humans, animals and the environment. This thought is at the core of the One Health concept, which aims to highlight the “synergistic benefit of closer cooperation between human, animal and environmental health sciences” [2]. One example of a benefit derived from the connection between humans and animals is animal-assisted interventions (AAIs). The term “AAI” has become an umbrella term in the human-animal interaction (HAI) field, referring to all interventions that incorporate some element of HAI to achieve the desired outcome [3,4]. Based on the definition presented by López-Cepero, in this review AAIs are defined as any intervention that incorporates an element of HAI with an unfamiliar animal, with the aim of improving a human health outcome [4]. Unfamiliar animals are defined as animals that are not owned by or living with participants. Most commonly AAIs use dogs as the intervention animal, but other animals such as cats, horses, birds or fish are also sometimes used [5,6].

Past research has predominantly focused on the benefits of AAIs for clinical populations, and has found generally beneficial effects [7,8]. Among autism and dementia patients, AAIs have been found to improve social interaction as well as reduce problematic behaviors such as aggression or agitation [9–12]. AAIs are especially beneficial for patients with mental disorders. Several systematic reviews have shown reductions of clinical symptoms of disorders such as anxiety, depression and schizophrenia after an AAI [6,13]. Simultaneously, AAIs have improved engagement and social interaction among patients with mental disorders [14]. In addition, AAIs have been shown to reduce stress and improve well-being among non-clinical populations, including the elderly, children and higher education students [6,15,16].

There is a particularly strong need for stress-reducing interventions among students at higher education institutions. A higher education institution is “any postsecondary institution of learning that usually affords, at the end of a course of study, a named degree, diploma, or certificate of higher studies“[17]. Due to a multitude of factors including navigating a new environment, a high academic workload and financial pressures, the prevalence of stress as well as symptoms of depression and anxiety disorders are worryingly high among higher education students worldwide (27). According to the Anxiety and Depression Association of America, for example, 85% of students feel overwhelmed by academic expectations and demands, 40% of students state that anxiety is a top concern, and 30% of students state that stress negatively affects their academic performance [19,20]. Similar results have been replicated among higher education students around the world [18,21,22]. The burden of mental health problems among students has been continuously increasing over the past years, and has been further exacerbated by decreased social contact and increased worries about health and finances during the COVID-19 pandemic [22,23].

In light of these findings, AAIs are becoming increasingly common at higher education institutions to improve and promote student mental health [24]. Such programs most commonly take the form of drop-in events where groups of students can freely interact with dogs and their handlers [25]. AAIs in higher education settings are low-cost and easily scalable, allowing them to reach a large proportion of the student body [24,26–28]. In addition, AAIs are not stigmatized like other traditional mental health services due to the overwhelmingly positive perception of AAIs among higher education students [24]. This makes AAIs an ideal universal intervention for mental health promotion efforts at higher education institutions [29,30]. To confidently implement AAIs in higher education settings, a comprehensive overview of the current state of research and good evidence on the effects of AAIs on the mental and cognitive outcomes of students is needed.

The objective of this systematic review was therefore to estimate the effects of AAIs implemented in higher education settings on (1) the mental health outcomes and (2) the cognitive outcomes of students. This review also aims to contribute evidence to the “shared medicines and interventions” subgroup of *The Lancet* One Health Commission.

## Methods

### Protocol and registration

A systematic review protocol was developed in keeping with the PRISMA-P 2015 statement [31]. This protocol was registered on PROSPERO on August 12^th^ 2020 with the registration number CRD42020196283 [32].

### Sources, search methods and eligibility criteria

The literature search was conducted from June 10^th^ to June 20^th^ 2020, and was designed to identify all published and unpublished experimental and observational trials on AAIs conducted in higher education settings. Medline/PubMed, PsycInfo, CINAHL, Web of Science, Embase, ERIC, and Scopus were searched. In addition, WALTHAM Science, HABRI Central and Animal and Society Institute as well as the database OpenGrey were searched. Reference lists from relevant systematic reviews and included studies were hand-searched for potentially relevant publications. Due to the large number of retrieved results, only randomized controlled trials (RCTs) that were published in a peer-reviewed journal were included in this review. Studies were included if they assessed the effect of an intervention using a living animal that was unfamiliar to participants as the sole intervention tool, for any mental health or cognitive outcome of higher education students. Mental health outcomes were considered those that describe a person’s emotional or psychological state [33], for example through self-perceived assessments of stress, anxiety or depression. We also included physiological outcome measures that reliably correlate with acute stress, such as blood pressure (BP), heart rate (HR) or cortisol levels [34]. By contrast, we considered cognitive outcomes those that describe a person’s cognitive functioning [35], for example through assessments of intelligence, concentration or attention. Specifically in the higher education context, we also considered cognitive outcomes to include academic outcomes such as test performance. Details on the eligibility criteria can be found in S1 Table, while details on the search strategy can be found in S2 File and S2 Table.

### Study selection

The selection process was conducted in two steps, using Covidence [36]. First, two independent reviewers (AH and EW) screened articles by title and abstract and voted on eligibility. Potential disagreements were resolved through regular discussions. Second, the full text of remaining articles was evaluated by both reviewers (AH and EW). If articles could not be found, the corresponding author was contacted. If there was no response within two weeks, the articles were excluded. Articles both reviewers agreed upon were included in the systematic review.

### Quality assessment

Only quantitative outcomes that were assessed by at least three studies and could thus be meaningfully combined in a quantitative synthesis were included in the quality assessment process. The risk of bias of the included studies was assessed independently by two reviewers (AH and EW), using the Cochrane Risk-of-Bias tool for Randomized Trials 2 (RoB 2) [37]. The version for individually randomized, parallel-group trials as well as the version for crossover trials were used [38,39].

### Data extraction

Data extraction was independently conducted by AH and EW using an Excel sheet. Data was collected on the study design, study participants, the intervention condition, the control condition, and reported outcomes. Conflicts were resolved through regular discussions. A full list of the extracted data items can be found in S3 File.

### Data synthesis

All studies were grouped according to the qualitative or quantitative outcomes they assessed. For stress, anxiety and depression, we further differentiated between chronic (long-term) and acute (short-term) outcomes. We defined acute outcomes as measuring how a person is feeling in a given moment, and chronic outcomes as measuring how a person is feeling over a longer period of time.

### Qualitative synthesis

Quantitative outcomes reported by less than three studies, as well as all qualitative outcomes, were summarized in a qualitative synthesis. Study results were briefly summarized for each outcome. Studies assessing mental health outcomes, physiological outcomes and cognitive outcomes were grouped together, and common trends in results were described.

### Quantitative synthesis

Quantitative outcomes reported by three or more studies were included in the quantitative synthesis. For both the meta-analyses and the albatross plots, potential multiplicity was eliminated by applying the following rules: First, if an outcome was reported across multiple time-points, the last reported measurement of the outcome which was not yet part of follow-up measurements was chosen. Second, if an outcome was reported using multiple measures and the reported measures were assumed to be interchangeable, only one of the included measures was chosen. This was the case in studies reporting both systolic and diastolic BP, where systolic BP was chosen, and in studies reporting both HF (high-frequency) and rMSSD (root mean square of successive differences) heart rate variability (HRV), where HF HRV was chosen.

To be included in a meta-analysis, studies needed to supply an effect size (Hedges’ g) of the post-test difference in mental health or cognitive outcomes between an intervention group and a control group, and had to be of good quality (rated as “low risk” or “some concerns” by the RoB 2). In addition, studies had to use comparable intervention and control conditions. Interventions generally fell into two categories: (1) interventions that allowed participants to freely interact with animals and their handlers (active intervention), and (2) interventions where an animal was present while participants’ primary focus was on a task (passive intervention). These tasks typically aimed to increase the stress levels of participants (stressors), such as timed math tasks. Interventions were additionally categorized based on the animal species used in the intervention condition. Control conditions broadly fell into four categories: (1) control groups that replaced the presence of an animal with a human (active human control), (2) control groups that replaced the presence of the animal with a different animal, a toy animal, or pictures/videos of an animal (active animal control), (3) control groups with an active component that was not a human or a different animal like yoga (active other control), and (4) control groups without any active component (no-treatment control). Meta-analyses were conducted for all outcomes where at least three studies reported an effect size, were of good quality, and used comparable intervention and control conditions. Due to the small number of studies included in each meta-analysis, it was not possible to conduct moderator analyses.

For eligible outcomes, meta-analyses were conducted using RStudio Version 1.3.959 [40]. Summary effect sizes as well as the corresponding 95% confidence interval (CI) were calculated using a random-effects model, and visualized using forest plots. The heterogeneity between included studies was assessed using the Q and I^2^ statistics. If Hedges’ g and its standard error (SE) was not reported in the original study, it was computed in RStudio Version 1.3.959, using the package “esc” [41]. Details of the conducted calculations can be found in S4 Table. Funnel plots were used to explore publication bias, and Egger’s test for funnel plot asymmetry was conducted.

Due to the limited number of studies included in the meta-analyses, albatross plots were used to extend the quantitative data synthesis. The albatross plot is a graphical tool that allows an approximation of effect sizes based on p-value and sample size. A detailed explanation of the albatross plot and the mathematical background can be found in the corresponding paper by Harrison et al. [42]. The eligibility criteria in place for the meta-analyses were not required for inclusion in the albatross plots. Albatross plots were created using Stata/SE 16.1 (Stata Statistical Software. College Station, TX: StataCorp LLC; 2019). Effect size contours were calculated based on the standardized mean difference (SMD). Contours corresponded to the effect sizes 0.2 (small effect), 0.5 (medium effect) and 0.8 (large effect). Since all included studies were randomized, an equal group size was assumed. As suggested by Harrison et al., if a p-value was presented as a threshold instead of an exact value (e.g. p<0.05), the threshold value was used as the exact value [42]. In addition, for any non-significant outcome without an exact p-value (e.g. p>0.05), a p-value of 1 was substituted [42]. If not reported in the original study, p-values were calculated by conducting unpaired two-sided Student’s t-tests in RStudio Version 1.3.959, using the command “t.test” and the mean, standard deviation and sample size provided [40]. If not otherwise specified in the study, a normal distribution of the data was assumed.

The threshold for statistical significance was set at p<0.05 for all conducted calculations. The code used for all calculations can be found under DOI: 10.6084/m9.figshare.19368047.

## Results

### Study selection

A complete search of all databases as described above yielded a total of 2.431 search results. Screening of the reference lists of included articles and identified systematic reviews contributed an additional 63 search results, giving a total of 2.494 identified results. Details on the exact number of results obtained from each database can be found in S2 Table. After removing duplicates and screening the articles by title and abstract, a total of 218 articles remained for full text screening. After the full text screening, 32 articles remained for inclusion in this systematic review. Of these 32 articles, three reported two separate eligible studies [43–45], bringing the total of individual studies included in this review to 35. Common reasons for study exclusion can be found in the PRISMA flow chart (Fig 1). Of the 35 studies, 30 were included in the quantitative data synthesis. After assessing eligibility, eight studies were included in the meta-analyses and 28 studies were included in the albatross plots.

**Fig 1:**
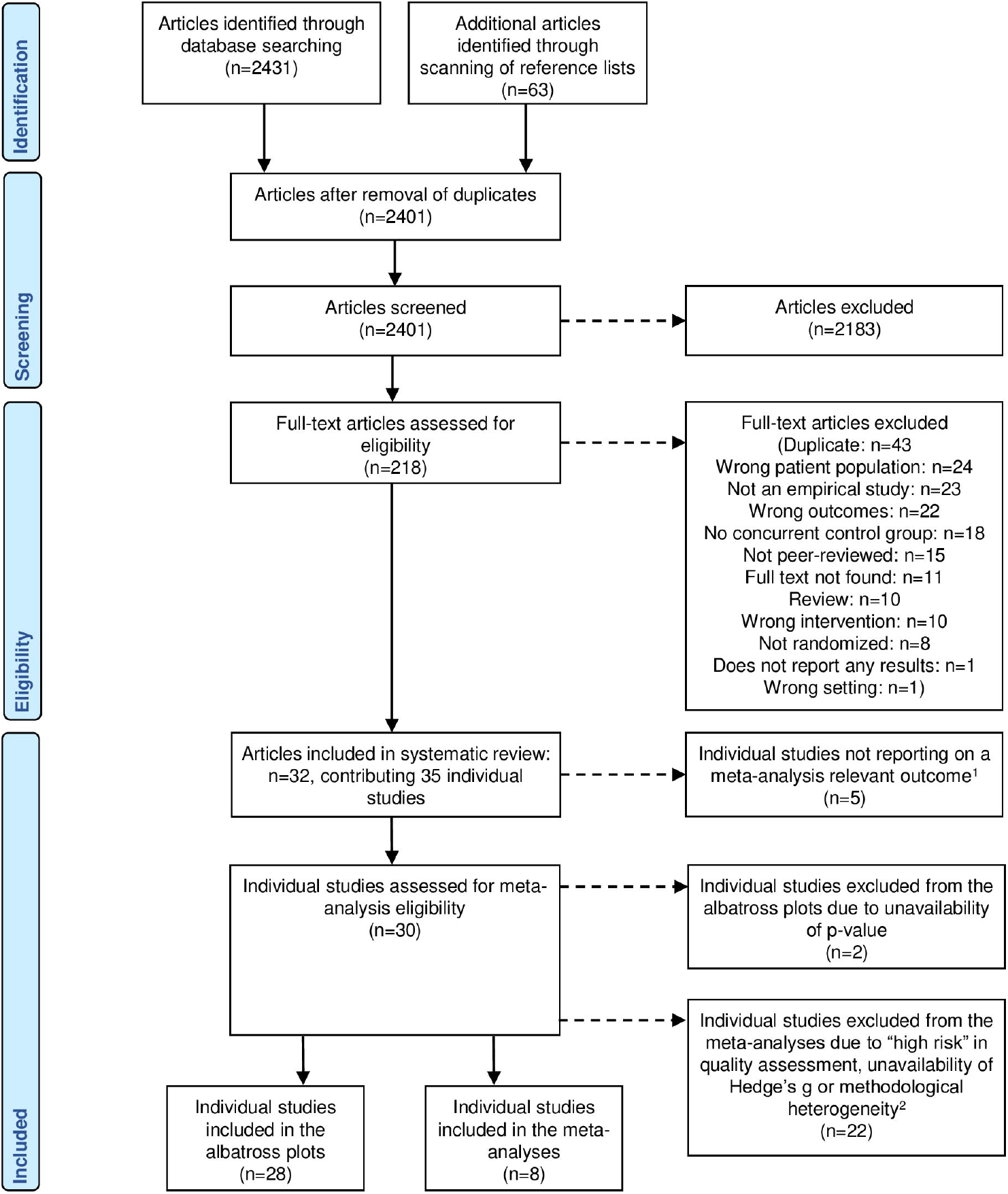
PRISMA flow chart. ^1^Quantitative outcomes assessed by less than three studies as well as qualitative outcomes were not eligible for inclusion in the meta-analyses. ^2^Methodological heterogeneity = heterogeneous for type of intervention condition (active/passive), animal used or type of control condition (active animal/active human/active other or no-treatment).

### Study characteristics

An overview of the most important extracted data items and study results can be found in the data extraction table (Table 1). Information on additional important study characteristics can be found in S5 Table. In general, most studies had more female than male participants, and participants were mostly of “typical” undergraduate age (mean 20.2 years, median 19.7 years). In almost all studies (n=29) the intervention animal was a dog [43,43,44,46–70]. Most studies (n=27) used active intervention conditions [43– 45,45,46,50,52–55,57,59–62,64–69,71–74], with most taking place in a group setting. In studies with active interventions, the animal-to-participant ratio was generally 1:3-5 participants. The remaining studies (n=8) [47–49,51,56,58,63,70] used passive intervention conditions that mostly took place in individual settings and included a stressor. In studies with passive interventions, the animal-to-participant ratio was generally 1:1. The most common control condition was a no-treatment control condition (n=27) [43,45,48–60,62– 67,69–72]. In most studies (n=28), intervention sessions took place only once per participant [43,43–45,45,47,48,50–58,60,64–66,68,70–74]. In general, intervention sessions were relatively short (mean 20.7 minutes, median 15 minutes).

**Table 1.**
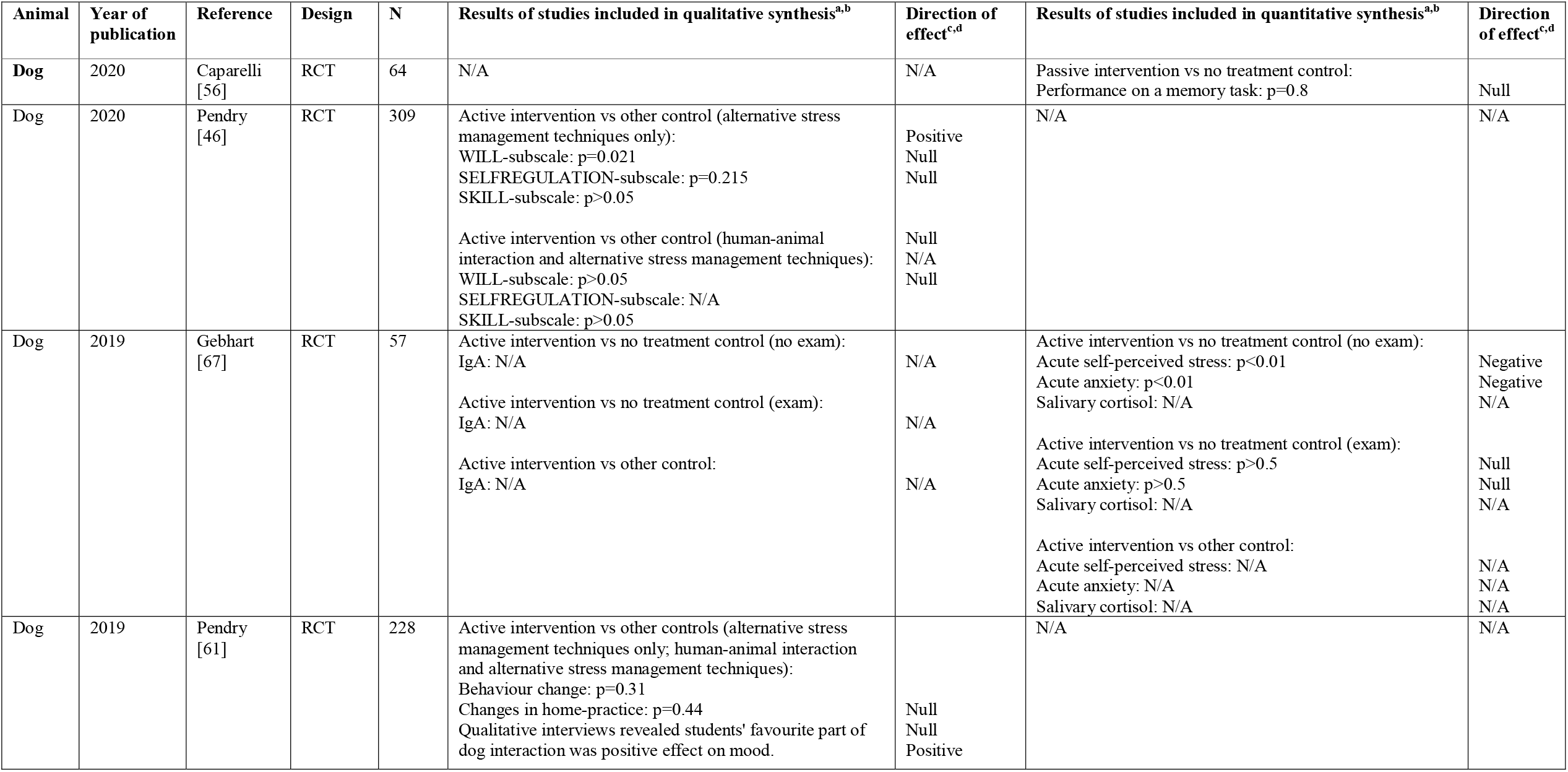
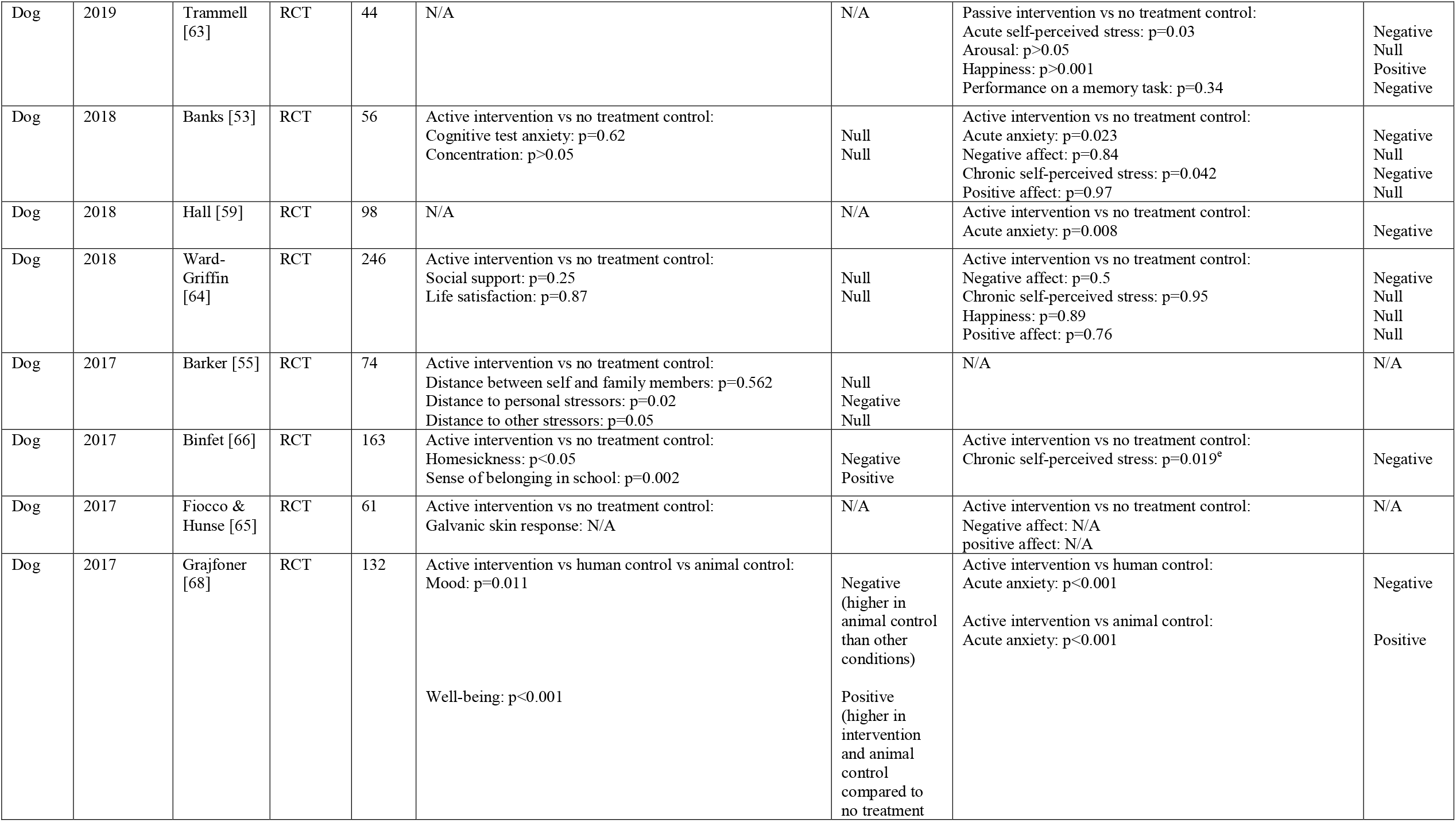
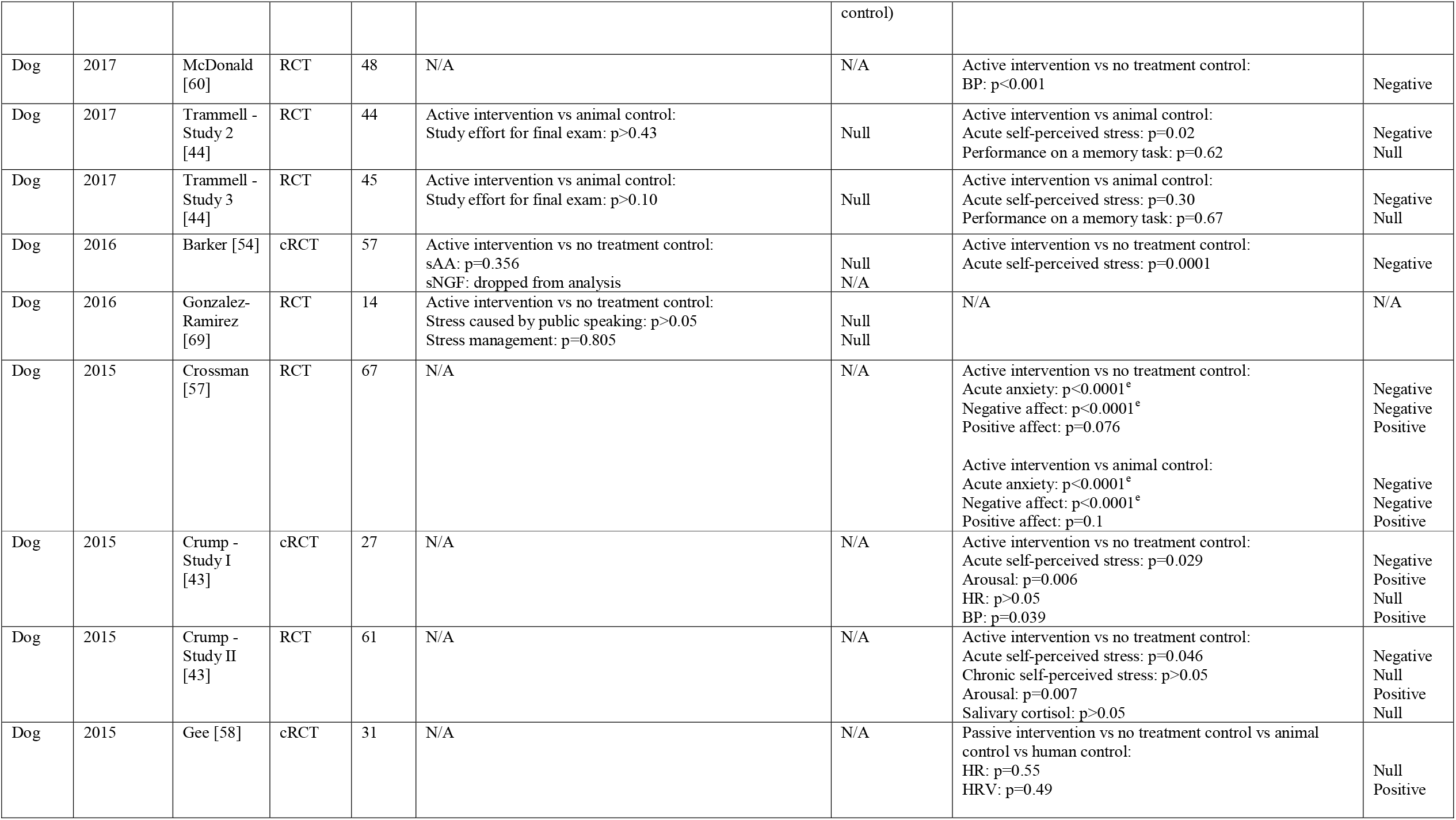
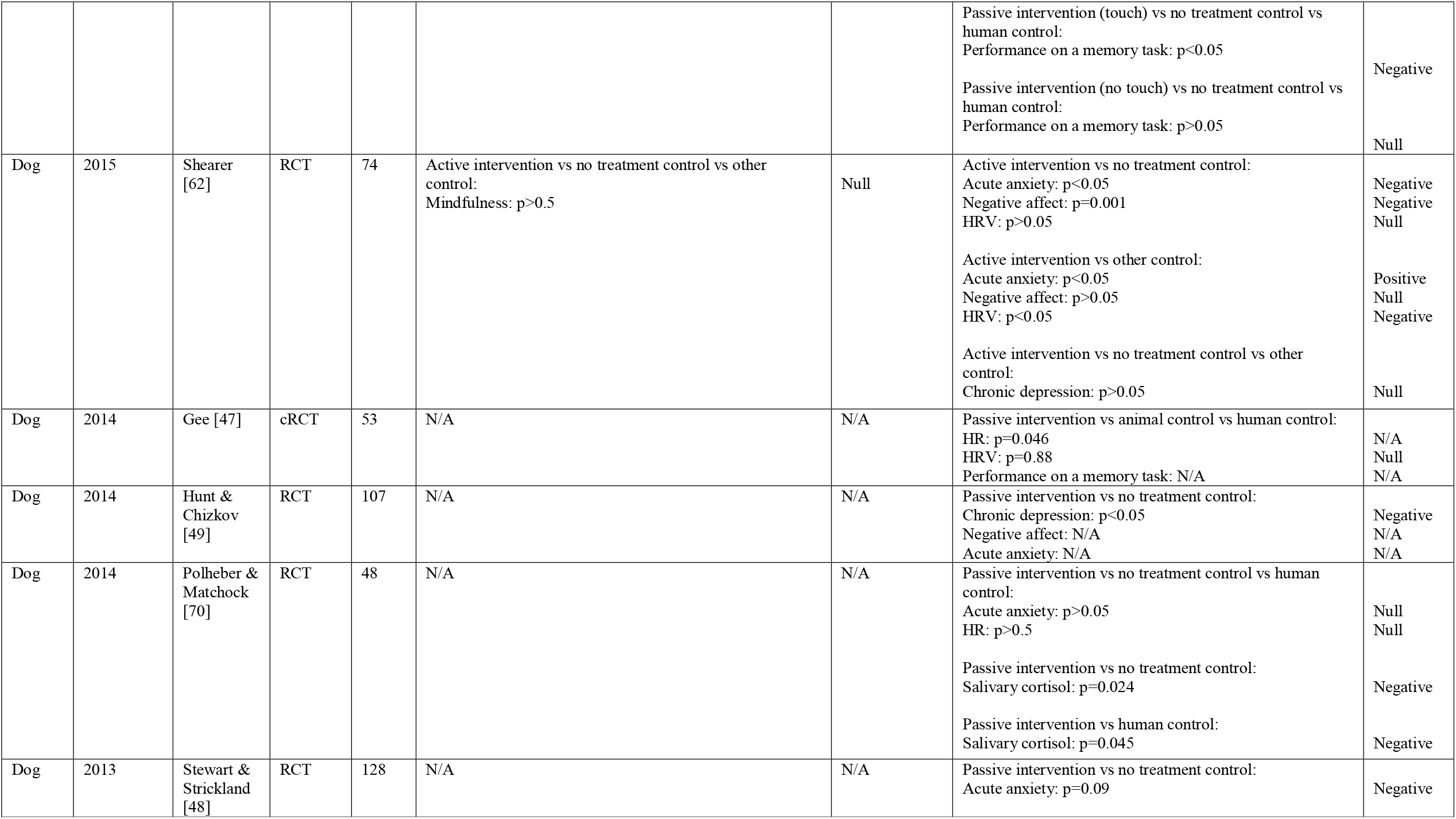
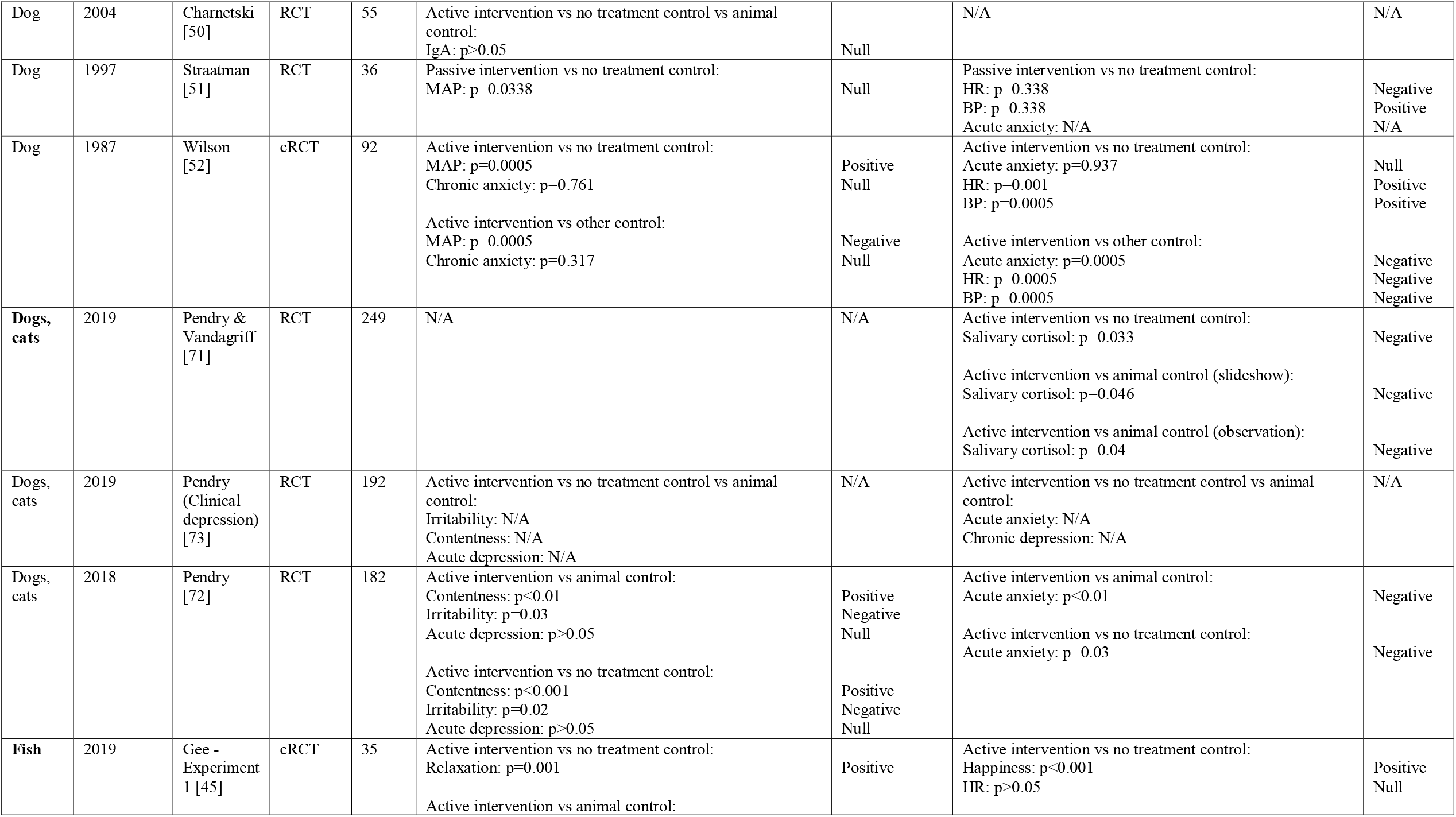
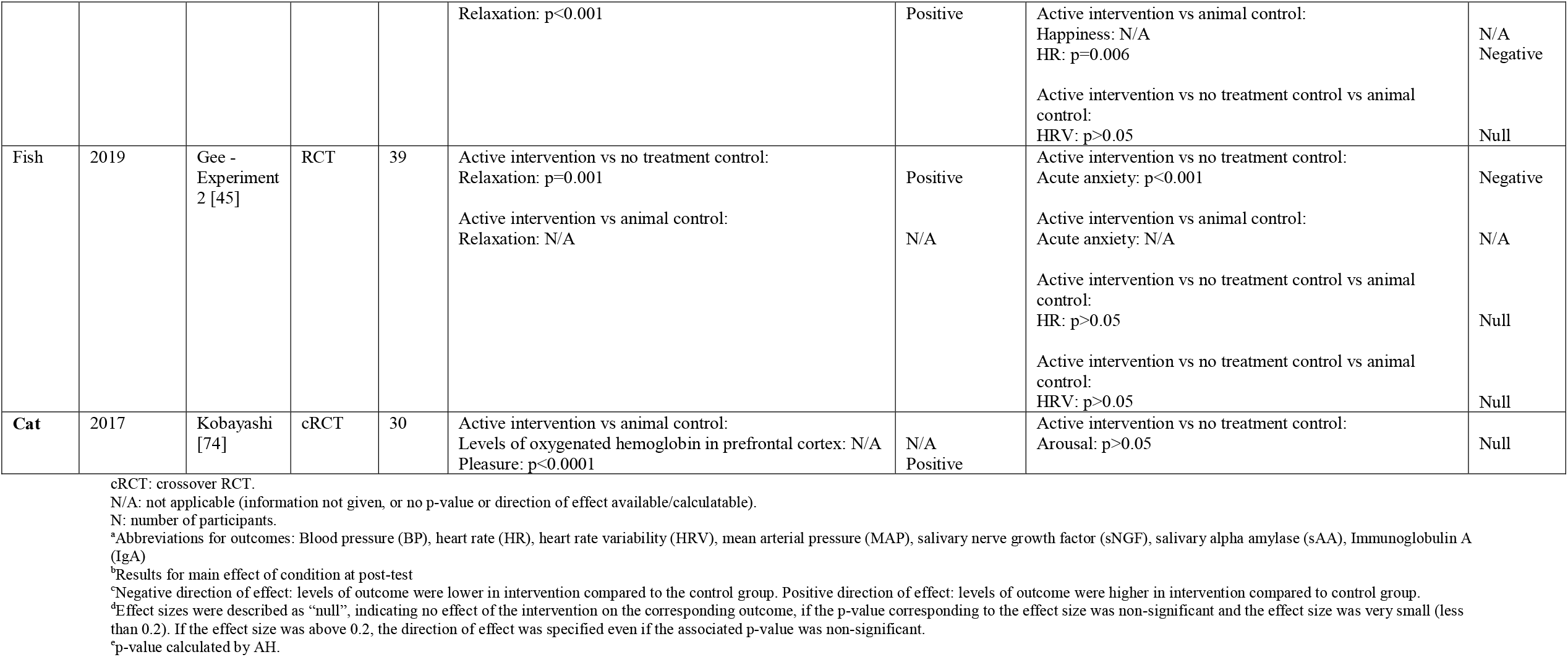
Data extraction table (n=35).

Outcomes were grouped into mental health outcomes, physiological outcomes and cognitive outcomes. Mental health outcomes were by far the most common (n=26) [43– 45,48,49,51–55,57,59,62–68,70,72–74], followed by physiological outcomes (n=14) [43,45,47,50,51,54,58,60,62,65,67,70,71,74] and cognitive outcomes (n=9) [44,46,47,53,56,58,61,63,69]. Most reported cognitive outcomes were related to students’ academic performance.

### Risk of bias within studies

Thirty studies were included in the quality assessment. Overall, 60 outcomes from 27 studies were classed as “some concerns” [43–45,47,48,51–54,56–58,60,62–68,70,71,73,74], 8 outcomes from 5 studies were classed as “high risk” [49,51,59,62,72], and no studies were classed as “low risk”. Common limitations included not reporting the method of allocation sequence generation or allocation sequence concealment. Additionally, blinding of participants and study personnel to a participants’ allocated condition was generally not possible due to the animal presence, although some studies tried to conceal the true study purpose from participants. Nonetheless, in most studies, both participants and study personnel were probably aware of their assigned condition, which may have affected self-reported outcomes. Finally, none of the included crossover RCTs gave information about potential carryover effects. An overview of quality assessment results for RCTs and crossover RCTs can be found in S6 and S7 Figs. Quality assessment results at the individual outcome level can be found in S6 and S7 Tables.

### Synthesis of results

#### Qualitative synthesis

Consistent with the study hypotheses, most studies reporting on negative mental health outcomes, including acute depression, homesickness and irritability, reported lower levels of these outcomes in the intervention group compared to the control group at post-test [52,66,72,73]. Only Wilson et al. did not report an effect of the intervention on chronic anxiety [52]. Similarly, some studies reporting on positive mental health outcomes reported higher levels of these outcomes in the intervention group compared to the control group at post-test [45,55,61,66,68,72–74]. However, a few studies also reported no effect of the intervention on positive mental health outcomes, including mood, life satisfaction and mindfulness [55,62,64,68]. Most studies reporting on physiological outcomes reported no effect of the intervention [50,51,54], although Fiocco & Hunse reported a smaller electrodermal response after a stressor in the intervention compared to the control group [65], and Wilson et al. found mean arterial pressure (MAP) to be significantly higher in the intervention compared to the control condition [52]. Similarly, most studies reporting on cognitive outcomes showed no effect of the intervention [46,53,61,69]. Only Pendry et al. found an improvement in test anxiety, attitude and study motivation [46] (Table 1).

#### Quantitative synthesis

The following outcomes were included in the quantitative synthesis: acute self-perceived stress, chronic self-perceived stress, negative affect, acute anxiety, arousal, chronic depression, happiness, positive affect, BP, HR, HRV, salivary cortisol, and performance on a memory task. Of these, meta-analyses were conducted for chronic self-perceived stress, negative affect, acute anxiety, positive affect and BP. All studies included in the meta-analyses used an active intervention condition, a dog as the intervention animal and a no-treatment control condition. The most important results are presented below. Detailed results for the remaining outcomes, including the albatross plots and the meta-analyses, can be found in S8 - S19 Figs.

Mental health outcomes were most common. For acute anxiety and self-perceived stress, most included studies showed a significant reduction at post-test. Acute anxiety was reported by 14 studies, of which four studies were combined in a meta-analysis [52,53,57,62]. The pooled Hedges’ g was -0.57 (95% CI -1.45, 0.31; Q=12.5, I^2=76%, p=0.006), indicating a medium-sized negative effect of the intervention (Fig 2). This result was mirrored by the albatross plot, where most studies clustered around the 0.5 to the 0.8 negative effect size contours (Fig 3). Acute self-perceived stress was reported by seven studies. Although not combinable in a meta-analysis, the albatross plot demonstrated that included studies showed a reduction of self-perceived stress with a medium to large effect size, with most results clustering around the 0.5 to the 0.8 negative effect size contours of the albatross plot (Fig 4).

**Fig 2.**
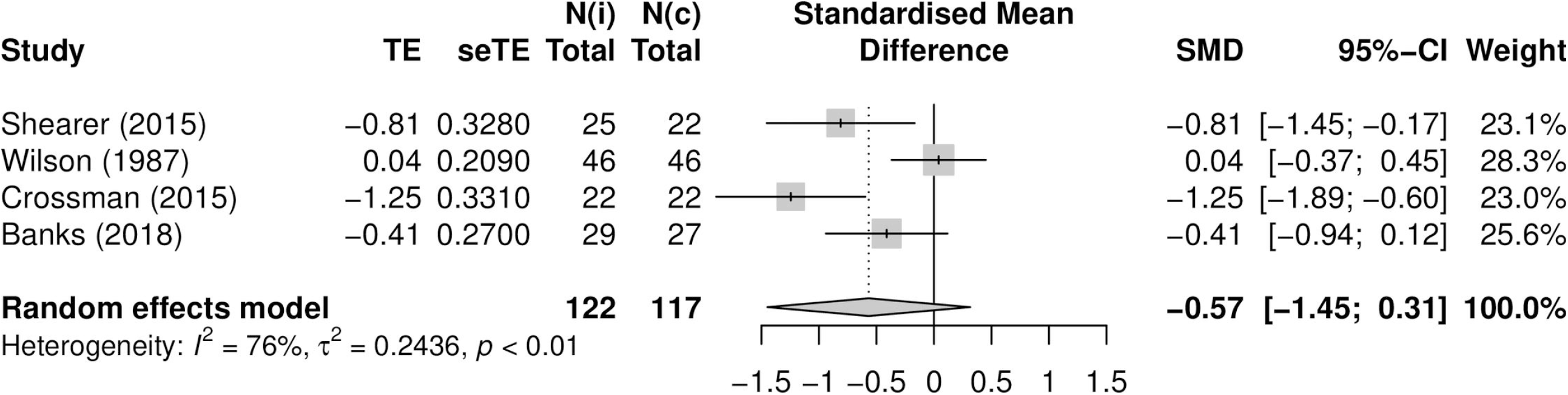
Forest plot acute anxiety (n=4). TE: Hedges’ g. seTE: standard error of Hedges’ g. N(i): number of participants in intervention condition. N(c): number of participants in control condition.

**Fig 3.**
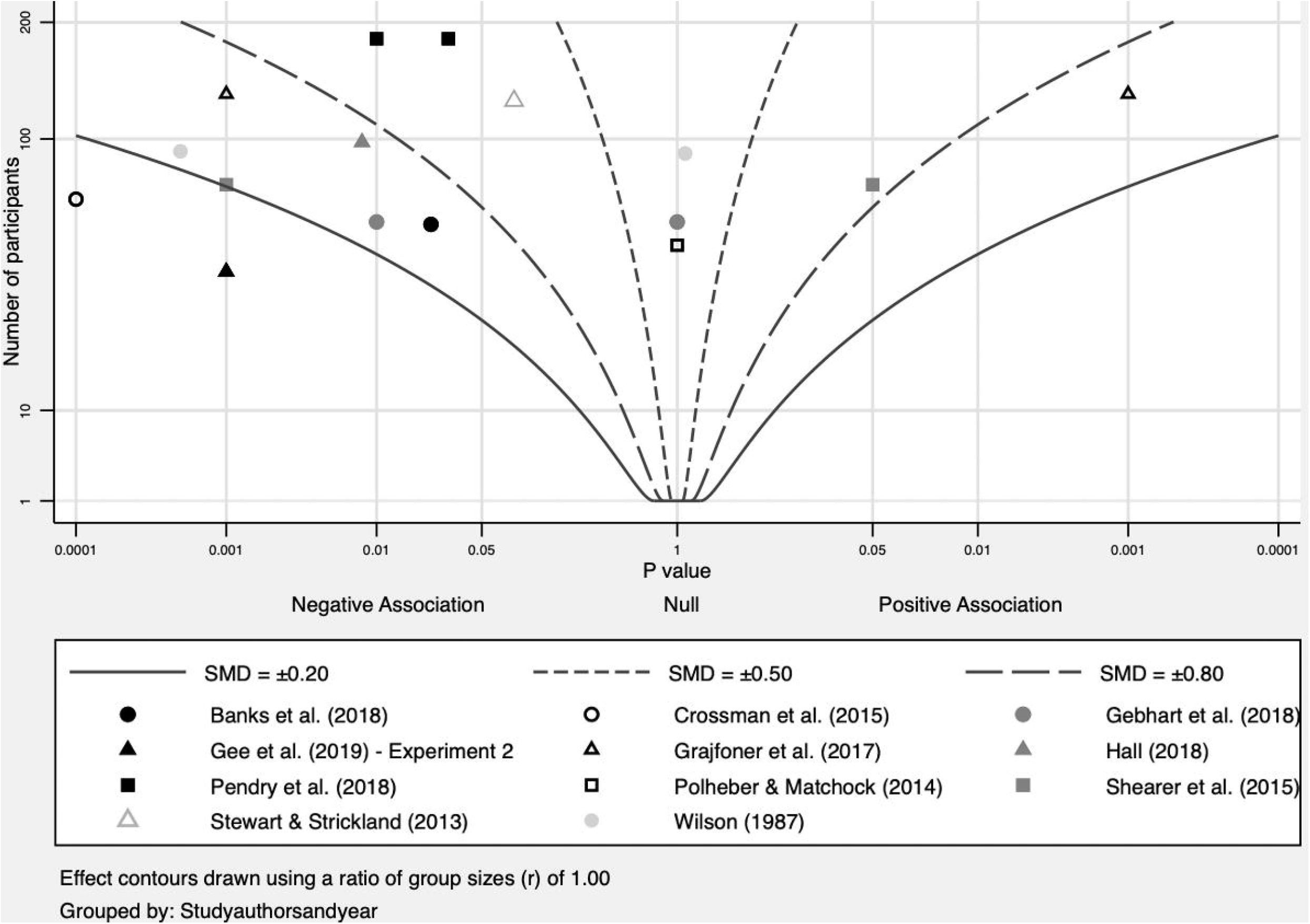
Albatross plot acute anxiety (n=11).

**Fig 4.**
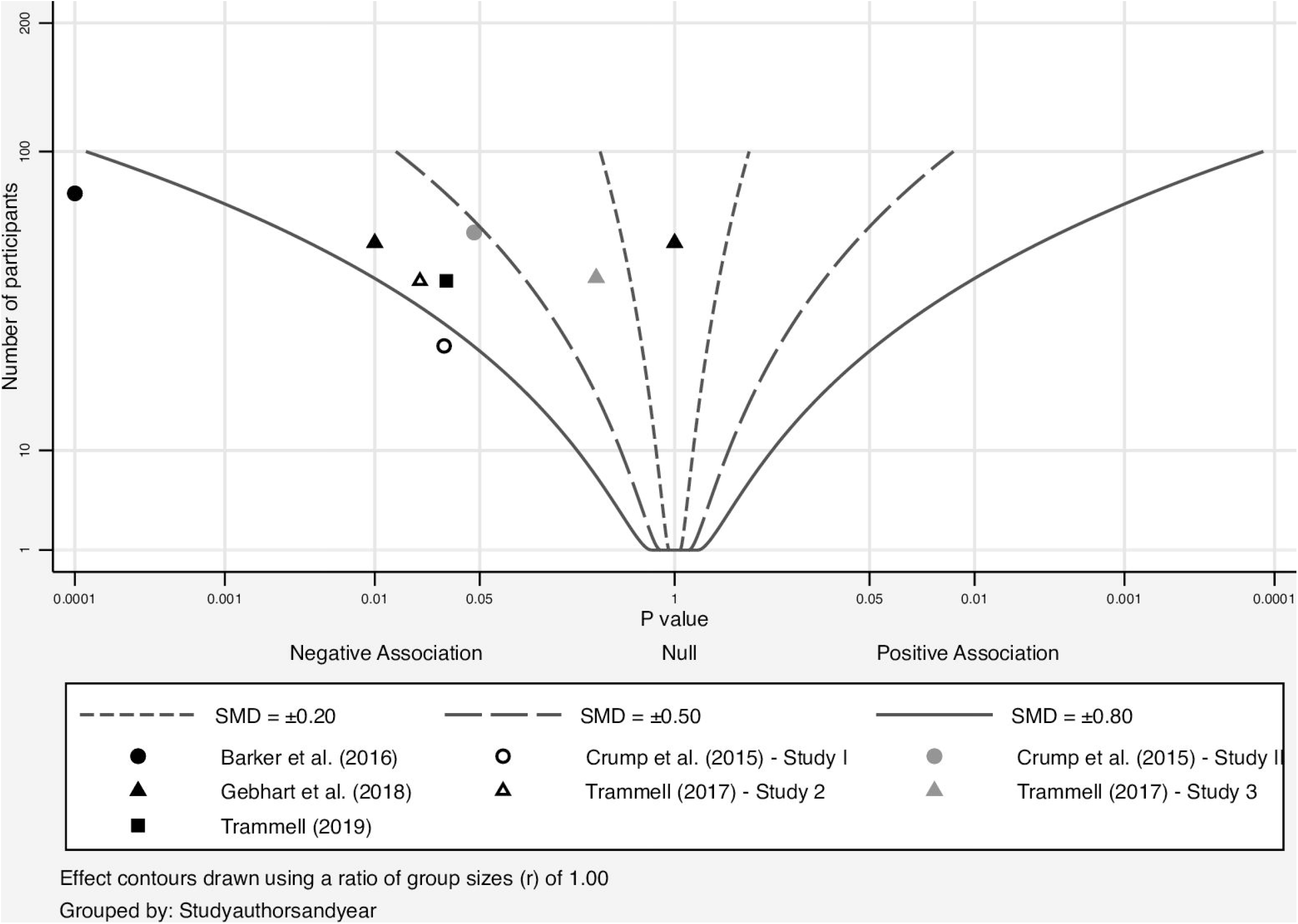
Albatross plot acute self-perceived stress (n=7).

Negative affect was reported by 6 studies, of which 4 studies were combined in a meta-analysis [53,57,62,64]. The pooled Hedges’ g was -0.47 (95% CI -1.46, 0.52; Q=15.3, I^2=80.4%, p=0.016), indicating a small- to medium-sized negative effect of the intervention (Fig 5). The albatross plot showed that while some studies showed a reduction of negative affect, other studies showed no effect (Fig 6).

**Fig 5.**
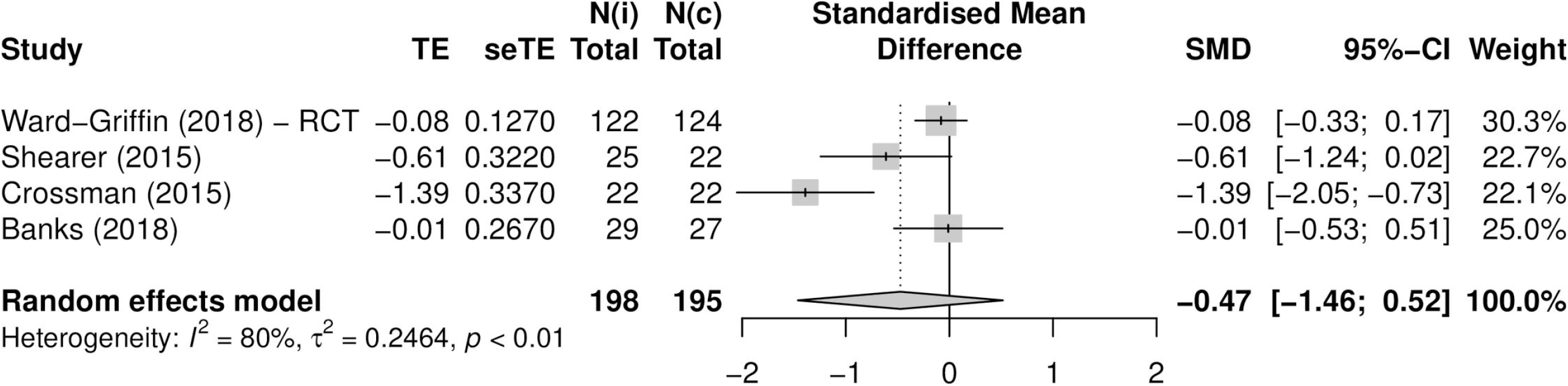
Forest plot negative affect (n=4). TE: Hedges’ g. seTE: standard error of Hedges’ g. N(i): number of participants in intervention condition. N(c): number of participants in control condition.

**Fig 6.**
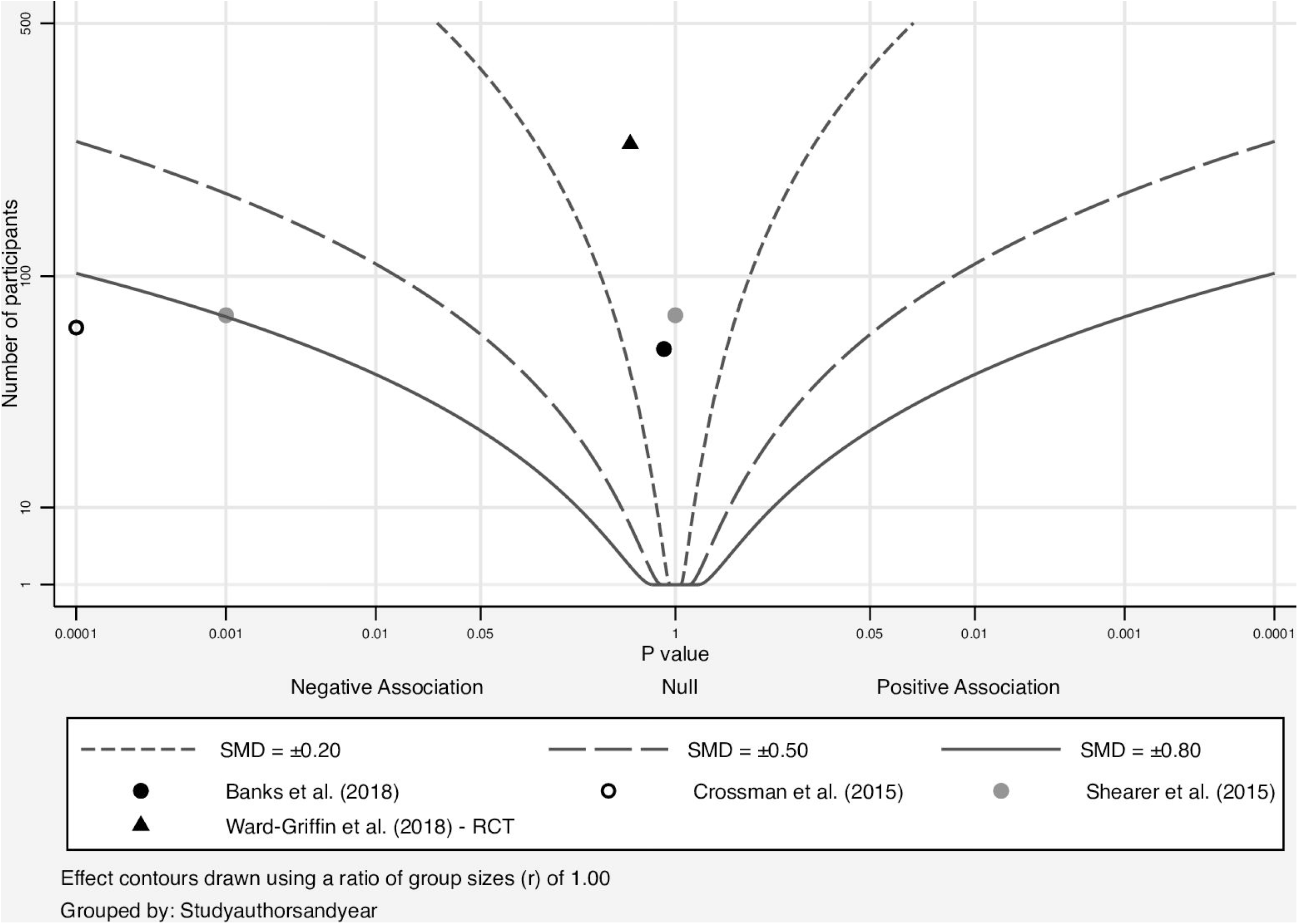
Albatross plot negative affect (n=5).

The tendency for some studies to show the expected effect while other studies showed no effect was also observed for the remaining mental health outcomes. Accordingly, a small negative effect of the intervention was observed for chronic self-perceived stress (pooled Hedges’ g: -0.23 (95% CI -0.57, 0.11; heterogeneity: Q=1.44, I^2=0%, p=0.49) and chronic depression, and a small positive effect was observed for positive affect (pooled Hedges’ g: 0.06 (95% CI -0.78, 0.90; heterogeneity: Q=3.97, I^2=49.6%, p=0.138), arousal and happiness. Forest plots and albatross plots for these outcomes can be found in S8 - S14 Figs.

Among the physiological outcomes, salivary cortisol was the only outcome to demonstrate the expected direction of effect. Salivary cortisol was reported by four studies. Although not combinable in a meta-analysis, included studies showed a small to medium negative effect on cortisol, with most results falling between the 0.3 and 0.8 effect size contours of the albatross plot. In contrast, among the 8 studies assessing HR, most included studies showed no effect on HR, with most results clustered around the middle of the albatross plot (Fig 7). This trend was mirrored by the studies assessing HRV.

**Fig 7.**
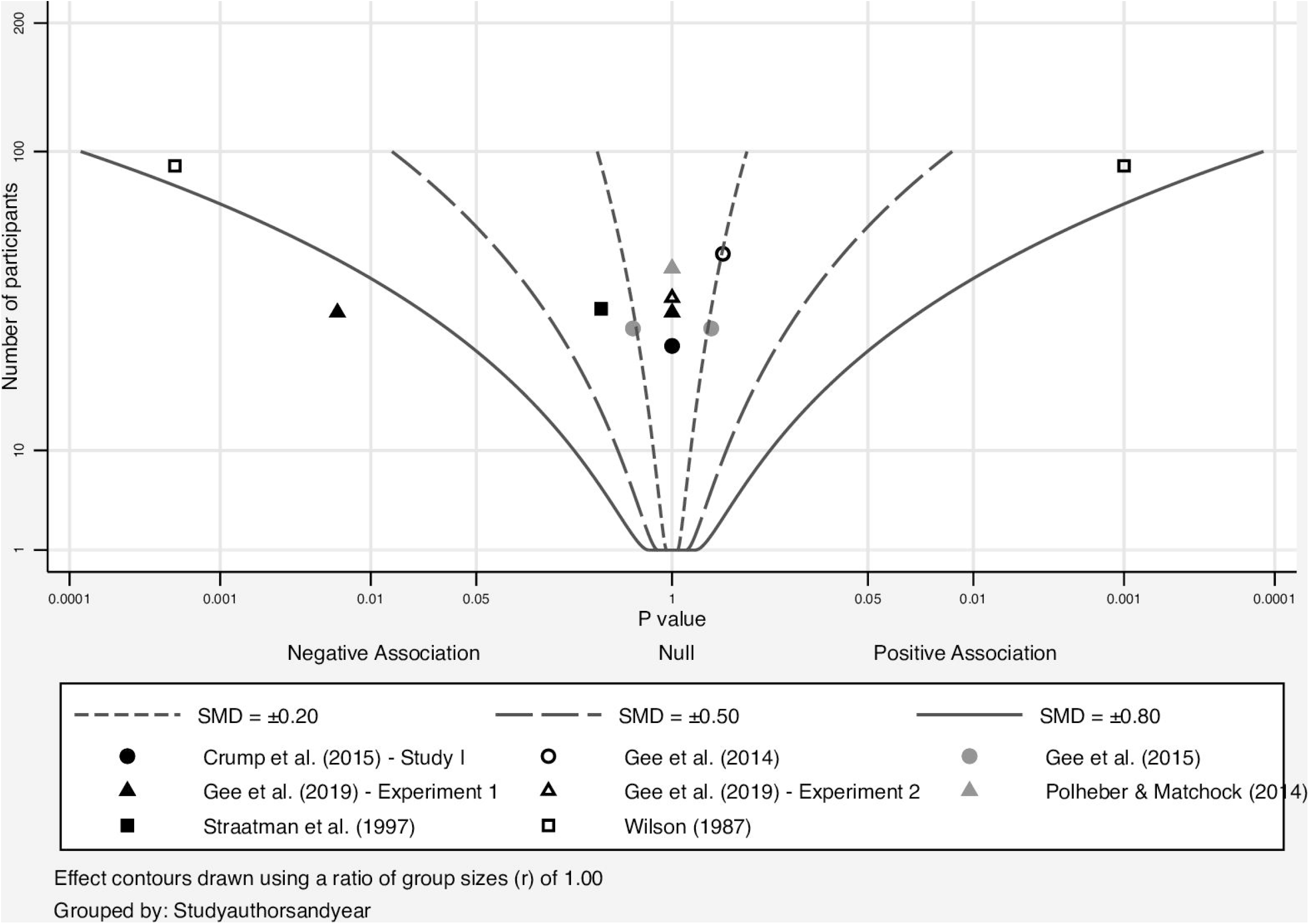
Albatross plot heart rate (n=8).

Interestingly, studies reporting on BP were very heterogeneous in terms of outcome. BP was reported by four studies, three of which were combinable in a meta-analysis. However, despite correcting for methodological heterogeneity, the results of studies included in the meta-analyses were very disparate in terms of both size and direction of effect. Additionally, the forest plot showed a very high, statistically significant level of heterogeneity between the included studies (Q=45.5, I^2=95.6%, p<0.0001). These levels of heterogeneity were significantly higher than for any other meta-analysis conducted. Accordingly, it was deemed inappropriate to statistically combine BP outcomes, and no pooled effect size was calculated. This strong heterogeneity was mirrored in the albatross plot, where results were spread out throughout the plot. Forest plots and albatross plots for physiological outcomes can be found in S15 - S18 Figs.

The only cognitive outcome included in the quantitative synthesis was performance on a memory task, reported by six studies. Overall, included studies suggested a very small negative effect of the intervention on memory task performance, with most results clustering around the 0.2 effect size contour of the albatross plot. The albatross plot can be found in S19 Fig.

### Risk of bias across studies

The funnel plot showed no evidence of publication bias, as confirmed by Egger’s regression test for funnel plot asymmetry (z= -1.74, p=0.081). The funnel plot can be found in S20 Fig.

## Discussion

### Summary of findings

The aim of this systematic review and meta-analysis was to assess the effect of AAIs implemented in higher education settings on the mental and cognitive outcomes of students. Additionally, we assessed the overall quality of included studies. In general, the results of this review suggest that AAIs in higher education settings are particularly effective at reducing acute feelings of anxiety and stress. The evidence is less clear for other mental health outcomes assessed in this review, but the included studies suggest a beneficial effect of AAIs on these outcomes as well. This review does not suggest a beneficial effect of AAIs on physiological or cognitive outcomes of students. Overall, the quality of included studies was moderate, with most studies being classed as “some concerns”.

### Mental health outcomes

The beneficial effects on acute mental health outcomes found in this review are in keeping with previous systematic reviews, which have shown AAIs to improve mental health outcomes in a large variety of populations. Several previous systematic reviews have shown significant reductions in self-perceived stress and anxiety in populations with and without pre-existing health conditions [5,8,15]. Similarly, previous reviews have shown AAIs to promote a positive mood, increase happiness and reduce depressive symptoms [6,8,75,76]. While the overall direction of effect of the included mental health outcomes was beneficial as expected, some studies reporting on mental health outcomes showed no effect of the intervention. Since formal moderator analyses were not possible in this review, we cannot say with certainty which, if any study characteristics are associated with this. The comparatively smaller effect sizes of chronic stress and depression, both assessed with instruments designed to detect changes in the mental state over longer periods of time, may point to limited long-term effects of AAIs, as suggested in previous literature [77–79].

Another possible contributor to differences in study results could be related to intervention design. Beetz et al. suggest that the beneficial effects of HAI could stem from an activation of the oxytocin (OT) system through sensory stimulation [8]. Specifically, they state that the closeness of the connection between human and animal, including the duration of the gaze from the animal as well as the presence and duration of physical contact with the animal, is an important factor in if and how much OT is released during HAI [8]. Since participants in studies using a passive intervention and stressors were not able to focus completely on the present animal and often did not even touch the animal in question, it is possible that not enough OT was released, thus explaining the lack of expected effects on mental health outcomes seen in some of these studies [48,49]. Since moderator analyses to confirm this hypothesis were not possible in this review, future research could explore whether the use of passive interventions and stressors is indeed associated with a reduced effect of AAIs on the mental health outcomes of higher education students.

### Physiological outcomes

Only three of the included studies provided results for cortisol, with two studies reporting reductions and one study reporting no effect on salivary cortisol at post-test [43,70,71]. This trend towards a reduction of cortisol at post-test is in keeping with other literature [8,80]. By contrast, studies assessing BP showed very mixed outcomes. This mixed effect of AAIs on BP has been reported in other systematic reviews, even though the overall trend seems to be that BP decreases post-AAI [5,8]. One possible explanation for the large discrepancies between BP results in this review and past reviews could be the poor reliability of BP as an outcome measure. Indeed, a study by Kelsey et al. showed that measures of cardiovascular reactivity, including BP, have a poor reliability across different typical stressor tasks [81]. BP can be affected by variables such as the posture of the participant, movement, respiration, sensory input or varying task demands [81]. All of these factors differed between the studies included in this review. Additionally, while all studies used a BP monitor, measurements were taken from different locations including the upper arm [43,52], the wrist [60] or the finger [51], which may also have contributed to the heterogeneity in results.

Interestingly, most included studies showed no effect of the intervention on HR or HRV. This is different from findings of other reviews, which have found an overall reduction of HR after an AAI in a variety of populations [5,82]. It is possible that AAIs may have less of an effect on physiological outcomes in a young, healthy population. Indeed, while Nimer & Lundahl found a significant improvement of physiological outcomes after an AAI, moderator analyses revealed that populations with disabilities showed significantly larger improvements than healthy populations did [16]. Additionally, it is possible that differences in effects between studies are again associated with intervention design: Most studies that assessed HR and HRV included a stressor in their intervention, thus likely triggering an acute stress response among participants. It is well established that, in response to an acute stressor, HR increases while HRV decreases [83]. Accordingly, it is possible that in studies with a stressor, the potential effect of an AAI on these physiological outcomes was not strong enough to compete with or alter the effects of the acute stress response. More studies without an incorporated stressor would be needed to judge the effects of AAIs on the physiological outcomes of students in a non-stressful situation.

### Cognitive outcomes

The studies included in this review showed no effect on the intervention on cognitive outcomes. This is an interesting finding, especially considering that past systematic reviews assessing the impact of AAIs on cognitive outcomes of children have found that the presence of animals to helps to create a productive learning environment [8,84]. Although these systematic reviews point out that there is little evidence that AAIs directly improve academic performance, they have nevertheless been shown to improve related cognitive outcomes like concentration, motivation, attention and social functioning [8,84]. However, Banks et al. hypothesized that while the presence of an animal may be beneficial for children, whose cognitive functions are still developing, there is less of an impact on these outcomes among higher education students, who are already at their peak of cognitive functioning [53]. The primary benefits of AAIs for this population therefore seem to be affective, not cognitive.

### Limitations of included evidence

Past systematic reviews in the AAI field have cited a limited availability of RCTs as a central limitation. However, our literature search yielded enough RCTs to answer our research questions. Of the 32 papers included in this review, 25 were published in 2015 or later, showing that this increase in RCT availability is quite a recent development. This is an encouraging finding, signaling the strong interest in this field from the scientific community. The quality of the RCTs included in this review was also judged to be satisfactory by the RoB2.

Nevertheless, some characteristics shared by the included studies may limit the generalizability of the results found in this review. First, participants in included studies were overwhelmingly female. This may be attributable to an increased interest in AAIs among females, as most studies recruited participants via self-selection, or to recruitment from traditionally majority-female degree programs, such as psychology or nursing (150,151). Since previous research has shown differences between males and females in, for example, responses to stressors, it is possible that the results may not be generalizable to both male and female students (152,153). Second, although the search strategy was designed to find publications using any intervention animal, almost all included studies used dogs. This may be due to the popularity of dogs as companion animals and the feelings of empathy and companionship associated with them, making them a popular choice for AAIs [85]. Additionally, dogs may have been the easiest option logistically since some studies cooperated with established university-based AAI programs that were already using dogs [44,46,53–55,61,68,71–73], and some studies used pet dogs of the researchers [71–73]. Nonetheless, it should be kept in mind that the results of this review represent the effects of AAIs using dogs and are likely less applicable to AAIs using other animals. Other limitations of included studies were small sample sizes, lack of sample size calculations and general lack of follow-up assessments.

### Limitations of the review process

The strength of this review lies in the use of the albatross plots to enrich the quantitative data synthesis, as well as the inclusion of RCTs only. Nonetheless some important limitations remain.

First, the strong methodological heterogeneity severely limited the comparability of included studies, as has been the case with many other reviews in the AAI field [5,6,13]. The heterogeneity also limited the number of studies included in the individual meta-analyses and limited our ability to conduct moderator analyses. This heterogeneity is at least partly attributable to the broadly defined eligibility criteria used in this review. Kazdin et al. have remarked that such broad eligibility criteria, where inclusion is based on the presence of an animal in the intervention as opposed to the proposed mechanism of the intervention, is one of the reasons for the methodological heterogeneity in most reviews in the AAI field [86]. This lack of a guiding theoretical framework in most reviews is exacerbated by the lack of an unanimously accepted theory on the mechanism of AAI effectiveness in the field [87]. In order to limit this issue in future research, systematic reviews should settle on a specific theoretical framework to guide their eligibility criteria in order to include only logically comparable studies [86].

Second, the albatross plots in this review were explicitly meant to allow a more inclusive overview of available data than what was available based on meta-analyses alone and were not meant to generate a usable summary statistic. The effect size contours superimposed on the plot are only approximations of the actual effect size (100). While they allow a visual interpretation of the general trend of the included studies in terms of effect size and direction, they are not exact and are not to be interpreted as such (100).

### Research gaps and implications

If possible, future reviews in this field could conduct moderator analyses to assess whether any study characteristics have an influence on study results, while future studies could focus on comparing different aspects of AAIs, perhaps by using multiple intervention conditions. For example, studies could explore whether incorporating a stressor in the study design or conducting an AAI in either a group or an individual setting influences the effect of AAIs on health outcomes.

Additionally, while a recent review suggested that AAI participation has no adverse effects for participating animals, research is limited and results remain conflicting [88]. There is even less research on potential benefits of AAI participation for animals [88]. Interestingly, research has suggested that following stress, trauma or abuse, animals can exhibit behavior similar to symptoms of human mental disorders such as depression or post-traumatic stress disorder [89,90]. Taking this into account, it is essential that the physical and mental health of animals participating in AAIs is protected. In the best case, AAIs should be mutually beneficial to animals and humans, thus making them a truly shared intervention in the spirit of One Health.

One of the goals of this review was to provide an evidence base that administrators at higher education institutions can use to decide whether to implement AAIs at their own campus. Despite the methodological limitations listed above, this review shows that AAIs can be effective in improving student mental health, especially acute feelings of anxiety and stress. Taking into consideration the high burden of mental health issues among students at higher education institutions, along with the unprecedented stress caused by the COVID-19 pandemic, higher education institutions will likely be facing an increasing demand for mental health support [29,91,92]. Due to their low cost, easy scalability and high popularity, AAIs present a good option for higher education institutions to improve student mental health [26].

This opportunity could be taken up particularly by universities outside of the US and Canada, where AAI programs are still rare. It has to be kept in mind, however, that while stress reduction efforts can certainly help, more structural changes should be implemented, which aim to reduce academic, social and financial pressures that impact students’ mental health. These could include, for example, an increased mental health budget at higher education institutions, reduced tuition fees and a mandatory salary for student internships [21,93,94].

### Conclusion

Overall, the results of this review suggest that AAIs in higher education settings can be effective at improving mental health outcomes of students and are particularly effective at reducing acute feelings of anxiety and stress. These findings have been replicated in many different settings and with a variety of populations. However, contrary to prior research, this review does not suggest a beneficial effect of AAIs on physiological or cognitive outcomes of students.

## Data Availability

All data relevant to the study is either contained within the Supporting Information files, or available in the public repository figshare (DOI: 10.6084/m9.figshare.19368047, URL: https://figshare.com/s/4f1e3a0de39bae34a2c0).

## Acknowledgements

The authors gratefully acknowledge Hélène Carabin for intellectual input at the beginning of the review process. Furthermore, we would like to thank Hilde Iren Flaatten, Medical Librarian at the University of Oslo, for her leading role in conducting the literature search.

## Supporting information

**S1 Table. Eligibility criteria**. ^a^We defined a parallel control group as a control group that experienced the control condition at the same time as the intervention group experienced the intervention condition. ^b^We defined randomization as true random allocation to either the intervention and control groups, or in the case of crossover studies, to the order of intervention and control groups.

**S2 File. Search strategy**.

**S2 Table. Overview of search results.**

**S3 File. List of extracted data items**.

**S4 Table. Calculations for meta-analysis**.

**S5 Table. Supplemental data extraction table**. n/s: not specified. ^a^Whether the intervention included a stressor (yes), no stressor (no) or took place shortly before exams. ^b^Whether the intervention took place in a group or individual format. ^c^Frequency sessions: whether sessions took place once (1) or more than once (>1).

**S6 Table. Quality assessment results for RCTs at the individual outcome level (n(outcomes)=51)**.

**S6 Figure. Overview of quality assessment results for RCTs (n(outcomes)=51)**. Results in %, absolute number of outcomes are inside the bars. Green corresponds to a rating of “low risk”, yellow to “some concerns”, and red to “high risk”.

**S7 Table. Quality assessment results for crossover RCTs at the individual outcome level (n(outcomes)=17)**.

**S7 Figure. Overview of quality assessment results for crossover RCTs (n(outcomes)=17)**. Results in %, absolute number of outcomes are inside the bars. Green corresponds to a rating of “low risk”, yellow to “some concerns”, and red to “high risk”.

**S8 Figure. Forest plot chronic self-perceived stress (n=3)**. TE: Hedges’ g. seTE: standard error of Hedges’ g. N(i): number of participants in intervention condition. N(c): number of participants in control condition.

**S9 Figure. Albatross plot chronic self-perceived stress (n=4).**

**S10 Figure. Albatross plot chronic depression (n=2)**.

**S11 Figure. Albatross plot arousal (n=4).**

**S12 Figure. Albatross plot happiness (n=3)**.

**S13 Figure. Forest plot positive affect (n=3)**. TE: Hedges’ g. seTE: standard error of Hedges’ g. N(i): number of participants in intervention condition. N(c): number of participants in control condition.

**S14 Figure. Albatross plot positive affect (n=3)**.

**S15 Figure. Albatross plot heart rate variability (n=5)**.

**S16 Figure. Albatross plot salivary cortisol (n=3)**.

**S17 Figure. Forest plot blood pressure (n=3)**. TE: Hedges’ g. seTE: standard error of Hedges’ g. N(i): number of participants in intervention condition. N(c): number of participants in control condition.

**S18 Figure. Albatross plot blood pressure (n=4)**.

**S19 Figure. Albatross plot performance on a memory task (n=5).**

**S20 Figure. Funnel plot (n=11)**.

**S21 Table. Completed PRISMA 2020 Checklist**.

## References

1. Andersen KG, Rambaut A, Lipkin WI, Holmes EC, Garry RF. The proximal origin of SARS-CoV-2. Nat Med. 2020 Apr;26(4):450–2.

2. Amuasi JH, Lucas T, Horton R, Winkler AS. Reconnecting for our future: The Lancet One Health Commission. The Lancet. 2020 May 9;395(10235):1469–71.

3. AVMA. Animal-assisted interventions: Definitions [Internet]. American Veterinary Medical Association. [cited 2020 May 4]. Available from: https://www.avma.org/policies/animal-assisted-interventions-definitions

4. López-Cepero J. Current Status of Animal-Assisted Interventions in Scientific Literature: A Critical Comment on Their Internal Validity. Animals. 2020 Jun;10(6):985.

5. Bert F, Gualano M, Camussi E, Pieve G, Voglino G, Siliquini R. Animal Assisted Intervention: a systematic review of benefits and risks. European Journal of Integrative Medicine. 2016 May 1;695–709.

6. Kamioka H, Okada S, Tsutani K, Park H, Okuizumi H, Handa S, et al. Effectiveness of animal-assisted therapy: A systematic review of randomized controlled trials. Complement Ther Med. 2014 Apr;22(2):371–90.

7. Maujean A, Pepping CA, Kendall E. A Systematic Review of Randomized Controlled Trials of Animal-Assisted Therapy on Psychosocial Outcomes. Anthrozoös. 2015 Mar 1;28(1):23–36.

8. Beetz A, Uvnäs-Moberg K, Julius H, Kotrschal K. Psychosocial and Psychophysiological Effects of Human-Animal Interactions: The Possible Role of Oxytocin. Front Psychol [Internet]. 2012 Jul 9 [cited 2021 Mar 9];3. Available from: https://www.ncbi.nlm.nih.gov/pmc/articles/PMC3408111/

9. Wood W, Fields B, Rose M, McLure M. Animal-Assisted Therapies and Dementia: A Systematic Mapping Review Using the Lived Environment Life Quality (LELQ) Model. Am J Occup Ther. 2017 Oct;71(5):7105190030p1–10.

10. Yakimicki ML, Edwards NE, Richards E, Beck AM. Animal-Assisted Intervention and Dementia: A Systematic Review. Clin Nurs Res. 2019;28(1):9–29.

11. O’Haire ME. Animal-assisted intervention for autism spectrum disorder: a systematic literature review. J Autism Dev Disord. 2013 Jul;43(7):1606–22.

12. Berry A, Borgi M, Francia N, Alleva E, Cirulli F. Use of assistance and therapy dogs for children with autism spectrum disorders: a critical review of the current evidence. J Altern Complement Med. 2013 Feb;19(2):73–80.

13. Brooks HL, Rushton K, Lovell K, Bee P, Walker L, Grant L, et al. The power of support from companion animals for people living with mental health problems: a systematic review and narrative synthesis of the evidence. BMC Psychiatry. 2018 05;18(1):31.

14. Jones MG, Rice SM, Cotton SM. Incorporating animal-assisted therapy in mental health treatments for adolescents: A systematic review of canine assisted psychotherapy. PLoS One [Internet]. 2019 Jan 17 [cited 2020 May 16];14(1). Available from: https://www.ncbi.nlm.nih.gov/pmc/articles/PMC6336278/

15. Ein N, Li L, Vickers K. The effect of pet therapy on the physiological and subjective stress response: A meta-analysis. Stress Health. 2018 Oct;34(4):477–89.

16. Nimer J, Lundahl B. Animal-assisted therapy: A meta-analysis. Anthrozoös. 2007;20(3):225–38.

17. Higher education [Internet]. Encyclopedia Britannica. [cited 2020 May 19]. Available from: https://www.britannica.com/topic/higher-education

18. Mortier P, Cuijpers P, Kiekens G, Auerbach RP, Demyttenaere K, Green JG, et al. The prevalence of suicidal thoughts and behaviours among college students: a meta-analysis. Psychol Med. 2018 Mar;48(4):554–65.

19. Facts Anxiety and Depression Association of America, ADAA [Internet]. [cited 2020 May 4]. Available from: https://adaa.org/finding-help/helping-others/college-students/facts

20. Austin E, Saklofske D, Mastoras S. Emotional intelligence, coping and exam-related stress in Canadian undergraduate students. Australian Journal of Psychology. 2010 Mar 1;62(1):42–50.

21. Bayram N, Bilgel N. The prevalence and socio-demographic correlations of depression, anxiety and stress among a group of university students. Soc Psychiatry Psychiatr Epidemiol. 2008 Aug;43(8):667–72.

22. Grützmacher J, Gusy B, Lesener T, Sudheimer S, Willige J. Gesundheit Studierender in Deutschland 2017. :168.

23. Campus Mental Health [Internet]. https://www.apa.org. [cited 2020 May 20]. Available from: https://www.apa.org/advocacy/higher-education/mental-health/index

24. Crossman MK, Kazdin AE. Chapter 24 - Animal Visitation Programs in Colleges and Universities: An Efficient Model for Reducing Student Stress. In: Fine AH, editor. Handbook on Animal-Assisted Therapy (Fourth Edition) [Internet]. San Diego: Academic Press; 2015 [cited 2020 Oct 3]. p. 333–7. Available from: http://www.sciencedirect.com/science/article/pii/B9780128012925000249

25. Gee NR, Fine AH, McCardle P. How Animals Help Students Learn: Research and Practice for Educators and Mental-Health Professionals. Taylor & Francis; 2017. 251 p.

26. Reynolds JA, Rabschutz L. Studying for Exams Just Got More Relaxing—Animal-Assisted Activities at the University of Connecticut Library. College & Undergraduate Libraries. 2011 Oct 1;18(4):359–67.

27. Wood E, Ohlsen S, Thompson J, Hulin J, Knowles L. The feasibility of brief dog-assisted therapy on university students stress levels: the PAwS study. J Ment Health. 2018 Jun;27(3):263–8.

28. Bell A. Paws for a Study Break: Running an Animal-Assisted Therapy Program at the Gerstein Science Information Centre. Partnership: The Canadian Journal of Library and Information Practice and Research [Internet]. 2013 Jun 1 [cited 2020 Sep 29];8(1). Available from: https://doaj.org

29. Vadivel R, Shoib S, El Halabi S, El Hayek S, Essam L, Gashi Bytyçi D, et al. Mental health in the post-COVID-19 era: challenges and the way forward. Gen Psychiatr. 2021;34(1):e100424.

30. Greenberg MT, Abenavoli R. Universal Interventions: Fully Exploring Their Impacts and Potential to Produce Population-Level Impacts. Journal of Research on Educational Effectiveness. 2017 Jan 2;10(1):40–67.

31. Moher D, Shamseer L, Clarke M, Ghersi D, Liberati A, Petticrew M, et al. Preferred reporting items for systematic review and meta-analysis protocols (PRISMA-P) 2015 statement. Systematic Reviews. 2015 Jan 1;4(1):1.

32. PROSPERO International Prospective Register of Systematic Reviews [Internet]. Animals in higher education settings: Do animal-assisted interventions improve physiological and subjective health outcomes of students? [cited 2020 Nov 15]. Available from: https://www.crd.york.ac.uk/prospero/display_record.php?RecordID=196283

33. WHO Mental health: a state of well-being [Internet]. WHO. World Health Organization; [cited 2020 May 30]. Available from: http://origin.who.int/features/factfiles/mental_health/en/

34. APA. Stress effects on the body [Internet]. https://www.apa.org. [cited 2020 Nov 15]. Available from: https://www.apa.org/topics/stress-body

35. Henderson C, Clements S, Corney S, Humin Y, Karmas R. Cognitive functioning: supporting people with mental health conditions [Internet]. Mental Health Coordinating Council (MHCC); 2015. Available from: https://www.mhcc.org.au/wp-content/uploads/2018/05/2016.02.17._supporting_cognitive_functioning-_mhcc_version_v._11__.pdf

36. Covidence - Better systematic review management [Internet]. [cited 2020 May 30]. Available from: https://www.covidence.org/home

37. Sterne JAC, Savović J, Page MJ, Elbers RG, Blencowe NS, Boutron I, et al. RoB 2: A revised tool for assessing risk of bias in randomised trials. BMJ. 2019;366:l4898.

38. Cochrane Methods. RoB 2.0 Guidance document for Parallel Trials [Internet]. Google Docs. [cited 2020 Dec 8]. Available from: https://drive.google.com/file/d/19R9savfPdCHC8XLz2iiMvL_71lPJERWK/view?usp=embed_facebook

39. Cochrane Methods. RoB 2.0 Guidance Document for Crossover Trials [Internet]. Google Docs. [cited 2020 Dec 8]. Available from: https://drive.google.com/file/d/18Ek-uW8HYQsUja8Lakp1yOhoFk0EMfPO/view?usp=embed_facebook

40. RStudio Team. RStudio: Integrated Development Environment for R. RStudio [Internet]. Boston, MA; 2020. Available from: http://www.rstudio.com/

41. Lüdecke D. esc: Effect Size Computation for Meta Analysis [Internet]. 2019 [cited 2021 Feb 25]. Available from: https://CRAN.R-project.org/package=esc

42. Harrison S, Jones HE, Martin RM, Lewis SJ, Higgins JPT. The albatross plot: A novel graphical tool for presenting results of diversely reported studies in a systematic review. Research Synthesis Methods. 2017;8(3):281–9.

43. Crump C, Derting TL. Effects of Pet Therapy on the Psychological and Physiological Stress Levels of First-Year Female Undergraduates. North American Journal of Psychology. 2015 Sep;17(3):575–90.

44. Trammell JP. The effect of therapy dogs on exam stress and memory. Anthrozoös. 2017;30(4):607–21.

45. Gee NR, Reed T, Whiting A, Friedmann E, Snellgrove D, Sloman KA. Observing Live Fish Improves Perceptions of Mood, Relaxation and Anxiety, But Does Not Consistently Alter Heart Rate or Heart Rate Variability. International Journal of Environmental Research and Public Health. 2019 Jan;16(17):3113.

46. Pendry P, Carr AM, Gee NR, Vandagriff JL. Randomized Trial Examining Effects of Animal Assisted Intervention and Stress Related Symptoms on College Students’ Learning and Study Skills. Int J Environ Res Public Health. 2020 15;17(6).

47. Gee NR, Friedmann E, Stendahl M, Fisk A, Coglitore V. Heart rate variability during a working memory task: Does touching a dog or person affect the response? Anthrozoös. 2014;27(4):513–28.

48. Stewart A, Strickland O. A Companion Animal in a Work Simulation: The Roles of Task Difficulty and Prior Companion-Animal Guardianship in State Anxiety. Society & Animals. 2013 Jan 1;21(3):249–65.

49. Hunt MG, Chizkov RR. Are Therapy Dogs Like Xanax? Does Animal-Assisted Therapy Impact Processes Relevant to Cognitive Behavioral Psychotherapy? Anthrozoös. 2014 Sep 1;27(3):457–69.

50. Charnetski CJ, Riggers S, Brennan FX. Effect of petting a dog on immune system function. Psychol Rep. 2004 Dec;95(3 Pt 2):1087–91.

51. Straatman I, Hanson EKS, Endenburg N, Mol JA. The Influence of a Dog on Male Students During a Stressor. Anthrozoös. 1997 Dec 2;10(4):191–7.

52. Wilson CC. Physiological responses of college students to a pet. Journal of Nervous and Mental Disease. 1987;175(10):606–12.

53. Banks J, McCoy C, Trzcinski C. Examining the Impact of a Brief Human-Canine Interaction on Stress and Attention. Human-Animal Interaction Bulletin. 2018 Jun 1;6(1):1–13.

54. Barker SB, Barker RT, McCain NL, Schubert CM. A Randomized Cross-over Exploratory Study of the Effect of Visiting Therapy Dogs on College Student Stress Before Final Exams. Anthrozoös. 2016 Jan 2;29(1):35–46.

55. Barker SB, Barker RT, McCain NL, Schubert CM. The Effect of a Canine-assisted Activity on College Student Perceptions of Family Supports and Current Stressors. Anthrozoös. 2017 Oct 2;30(4):595–606.

56. Capparelli AL, Miller QC, Wright DB, London K. Canine-Assisted Interviews Bolster Informativeness for Negative Autobiographical Memories. Psychol Rep. 2020 Feb;123(1):159–78.

57. Crossman MK, Kazdin AE, Knudson K. Brief unstructured interaction with a dog reduces distress. Anthrozoös. 2015;28(4):649–59.

58. Gee NR, Friedmann E, Coglitore V, Fisk A, Stendahl M. Does Physical Contact with a Dog or Person Affect Performance of a Working Memory Task? Anthrozoös. 2015 Sep 2;28(3):483–500.

59. Hall D. Nursing Campus Therapy Dog: A Pilot Study. Teaching and Learning in Nursing. 2018 Oct 1;13(4):202–6.

60. McDonald S, McDonald E, Roberts A. Effects of novel dog exposure on college students’ stress prior to examination. North American Journal of Psychology. 2017;19(2):477–84.

61. Pendry P, Kuzara S, Gee NR. Evaluation of Undergraduate Students’ Responsiveness to a 4-Week University-Based Animal-Assisted Stress Prevention Program. Int J Environ Res Public Health. 2019 Sep 10;16(18).

62. Shearer A, Hunt M, Chowdhury M, Nicol L. Effects of a brief mindfulness meditation intervention on student stress and heart rate variability. International Journal of Stress Management. 2016;23(2):232–54.

63. Trammell J. Therapy Dogs Improve Student Affect but Not Memory. Anthrozoös. 2019 Sep 3;32:691–9.

64. Ward-Griffin E, Klaiber P, Collins HK, Owens RL, Coren S, Chen FS. Petting away pre-exam stress: The effect of therapy dog sessions on student well-being. Stress Health. 2018 Aug;34(3):468–73.

65. Fiocco AJ, Hunse AM. The Buffer Effect of Therapy Dog Exposure on Stress Reactivity in Undergraduate Students. Int J Environ Res Public Health [Internet]. 2017 Jul [cited 2020 Dec 15];14(7). Available from: https://www.ncbi.nlm.nih.gov/pmc/articles/PMC5551145/

66. Binfet J-T. The Effects of Group-Administered Canine Therapy on University Students’ Wellbeing: A Randomized Controlled Trial. Anthrozoös. 2017 Jul 3;30(3):397–414.

67. Gebhart V, Buchberger W, Klotz I, Neururer S, Rungg C, Tucek G, et al. Distraction-focused interventions on examination stress in nursing students: Effects on psychological stress and biomarker levels. A randomized controlled trial. Int J Nurs Pract. 2020 Feb;26(1):e12788.

68. Grajfoner D, Harte E, Potter LM, McGuigan N. The Effect of Dog-Assisted Intervention on Student Well-Being, Mood, and Anxiety. Int J Environ Res Public Health [Internet]. 2017 May [cited 2020 Dec 15];14(5). Available from: https://www.ncbi.nlm.nih.gov/pmc/articles/PMC5451934/

69. González-Ramírez MT, Landaverde-Molina ÓD, Morales-Rodríguez D, Landero-Hernández R. Taller de manejo de ansiedad a hablar en público con la participación de perros de terapia. Ansiedad y Estrés. 2016 Jan 1;22(1):5–10.

70. Polheber JP, Matchock RL. The presence of a dog attenuates cortisol and heart rate in the Trier Social Stress Test compared to human friends. J Behav Med. 2014 Oct;37(5):860–7.

71. Pendry P, Vandagriff JL. Animal Visitation Program (AVP) Reduces Cortisol Levels of University Students: A Randomized Controlled Trial. AERA Open. 2019 Apr 1;5(2):2332858419852592.

72. Pendry P, Carr A, Roeter S, Vandagriff J. Experimental trial demonstrates effects of animal-assisted stress prevention program on college students’ positive and negative emotion. 2018 Mar 1;81–97.

73. Pendry P, Vandagriff J, Carr A. Clinical depression moderates effects of animal-assisted stress prevention program on college students’ emotion. Journal of Public Mental Health. 2019 May 31;18.

74. Kobayashi A, Yamaguchi Y, Ohtani N, Ohta M. The effects of touching and stroking a cat on the inferior frontal gyrus in people. Anthrozoös. 2017;30(3):473–86.

75. Morrison M. Health Benefits of Animal-Assisted Interventions. Complementary Health Practice Review. 2007 Jan 1;12:51–62.

76. Souter MA, Miller MD. Do animal-assisted activities effectively treat depression: a meta-analysis [Internet]. Database of Abstracts of Reviews of Effects (DARE): Quality-assessed Reviews [Internet]. Centre for Reviews and Dissemination (UK); 2007 [cited 2020 May 4]. Available from: https://www.ncbi.nlm.nih.gov/books/NBK74080/

77. Hillen N. Animal-assisted intervention on college campuses. TCNJ Journal of Student Scholarship. 2020;22:11.

78. Stern C, Chur-Hansen A. Methodological Considerations in Designing and Evaluating Animal-Assisted Interventions. Animals. 2013 Mar;3(1):127–41.

79. Serpell J, McCune S, Gee N, Griffin JA. Current challenges to research on animal-assisted interventions. Applied Developmental Science. 2017 Jul 3;21(3):223–33.

80. Lundqvist M, Carlsson P, Sjödahl R, Theodorsson E, Levin L-Å. Patient benefit of dog-assisted interventions in health care: a systematic review. BMC Complement Altern Med. 2017 Jul 10;17(1):358.

81. Kelsey RM, Ornduff SR, Alpert BS. Reliability of cardiovascular reactivity to stress: internal consistency. Psychophysiology. 2007 Mar;44(2):216–25.

82. Ein N, Li L, Vickers K. The effect of pet therapy on the physiological and subjective stress response: A meta-analysis. Stress and Health. 2018 Jun 8;34.

83. Chu B, Marwaha K, Sanvictores T, Ayers D. Physiology, Stress Reaction. In: StatPearls [Internet]. Treasure Island (FL): StatPearls Publishing; 2021 [cited 2021 Apr 21]. Available from: http://www.ncbi.nlm.nih.gov/books/NBK541120/

84. Brelsford VL, Meints K, Gee NR, Pfeffer K. Animal-Assisted Interventions in the Classroom—A Systematic Review. Int J Environ Res Public Health [Internet]. 2017 Jul [cited 2020 May 4];14(7). Available from: https://www.ncbi.nlm.nih.gov/pmc/articles/PMC5551107/

85. Custance D, Mayer J. Empathic-like responding by domestic dogs (Canis familiaris) to distress in humans: an exploratory study. Anim Cogn. 2012 Sep 1;15(5):851–9.

86. Kazdin AE. Strategies to improve the evidence base of animal-assisted interventions. Applied Developmental Science. 2017 Apr 3;21(2):150–64.

87. Borrego J, Rodríguez-Franco L, Perea-Mediavilla M, Blanco N, Tejada A, Blanco A. Animal-assisted Interventions: Review of Current Status and Future Challenges. International Journal of Psychology and Psychological Therapy. 2014 Mar 1;14:85–101.

88. Glenk LM. Current Perspectives on Therapy Dog Welfare in Animal-Assisted Interventions. Animals. 2017 Feb;7(2):7.

89. Ferdowsian HR, Durham DL, Kimwele C, Kranendonk G, Otali E, Akugizibwe T, et al. Signs of Mood and Anxiety Disorders in Chimpanzees. PLOS ONE. 2011 Jun 16;6(6):e19855.

90. PTSD in Dogs [Internet]. CVMBS News. 2010 [cited 2021 Apr 22]. Available from: https://vetmed.tamu.edu/news/pet-talk/ptsd-in-dogs/

91. Son C, Hegde S, Smith A, Wang X, Sasangohar F. Effects of COVID-19 on College Students’ Mental Health in the United States: Interview Survey Study. J Med Internet Res [Internet]. 2020 Sep 3 [cited 2021 Mar 9];22(9). Available from: https://www.ncbi.nlm.nih.gov/pmc/articles/PMC7473764/

92. Cao W, Fang Z, Hou G, Han M, Xu X, Dong J, et al. The psychological impact of the COVID-19 epidemic on college students in China. Psychiatry Research. 2020 May 1;287:112934.

93. Heckman S, Lim H, Montalto C. Factors Related to Financial Stress among College Students. Journal of Financial Therapy. 2014 Aug 1;5.

94. Hamaideh S. Stressors and Reactions to Stressors Among University Students. The International journal of social psychiatry. 2011 Jan 20;57:69–80.

