## Supplementary material for "Animals in higher education settings: Do animal-assisted interventions improve mental and cognitive health outcomes of students? A systematic review and meta-analysis": S1_Table

| **PICOS** criteria | Eligibility criteria |
|---|---|
| **P**opulation | • Participants are students of a higher education institution<br>• Participants can be with or without pre-existing health conditions |
| **I**ntervention | • Using a live animal<br>• Using an animal that is unfamiliar to participants<br>• Animal is the sole intervention tool<br>• Intervention takes place in a higher education setting<br>• Aim of the intervention is to improve any mental health or cognitive outcome of students |
| **C**ontrol | • Presence of a parallel control group[a] |
| **O**utcome | • Any mental health outcomes<br>• Any cognitive outcomes |
| **S**tudy design | • Study design must be RCT or crossover RCT with a parallel control group<br>• Study design must include true randomization[b]<br>• Published in a peer-reviewed journal |
