## Supplementary material for "Animals in higher education settings: Do animal-assisted interventions improve mental and cognitive health outcomes of students? A systematic review and meta-analysis": S2_File

**Database: Medline (Ovid)**
**Ovid MEDLINE(R) ALL** 1946 to June 09, 2020
Date searched: 10.06.2020
Number of hits: 307
Search Strategy:
1 animal assisted therapy/ or equine-assisted therapy/ or hippotherapy.tw,kf.
2 ((dog* or cat or cats or horse* or equine* or canine* or dolphin* or guinea pig* or rat or rats or rabbit* or chinchilla* or hamster*) adj1 (assisted or facilitated or visitation*)).tw,kf.
3 ((animal* or dog* or cat or cats or horse* or equine* or canine* or pet or pets or dolphin* or guinea pig* or rat or rats or rabbit* or chinchilla* or hamster*) adj1 therap*).tw,kf.
4 (animal assisted or animal facilitated or animal visitation* or pet-facilitated or pet assisted or pet visitation*).tw,kf.
5 (service adj (animal* or dog* or cat or cats or horse* or equine* or canine* or pet or pets or dolphin* or guinea pig* or rat or rats or rabbit* or chinchilla* or hamster*)).tw,kf.
6 ((human or people) adj1 (animal* or dog* or cat or cats or horse* or equine* or canine* or pet or pets or dolphin* or guinea pig* or rat or rats or rabbit* or chinchilla* or hamster*) adj1 (interaction* or activit* or therap* or intervention*)).tw,kf.
7 or/1-6
8 (school* or student* or classroom* or universit* or college* or education* or curricul* or campus* or diploma* or baccalaureate or higher stud* or graduat* or undergraduat* or facult* or learning environment or academic*).tw,kf.
9 exp Students/ or exp education/ or curriculum/ or exp schools/
10 or/8-9
11 7 and 10

**Database: Embase (Ovid)**
**Embase Classic+Embase 1947 to 2020 June 09**
Date searched: 10.06.2020
Number of hits: 378
Number of hits conference abstracts: 162
Search Strategy:
1 animal assisted therapy/ or hippotherapy/ or pet therapy/ or hippotherapy.tw,kw.
2 ((dog* or cat or cats or horse* or equine* or canine* or dolphin* or guinea pig* or rat or rats or rabbit* or chinchilla* or hamster*) adj1 (assisted or facilitated or visitation*)).tw,kw.
3 ((animal* or dog* or cat or cats or horse* or equine* or canine* or pet or pets or dolphin* or guinea pig* or rat or rats or rabbit* or chinchilla* or hamster*) adj1 therap*).tw,kw.
4 (animal assisted or animal facilitated or animal visitation* or pet-facilitated or pet assisted or pet visitation*).tw,kw.
5 (service adj (animal* or dog* or cat or cats or horse* or equine* or canine* or pet or pets or dolphin* or guinea pig* or rat or rats or rabbit* or chinchilla* or hamster*)).tw,kw.
6 ((human or people) adj1 (animal* or dog* or cat or cats or horse* or equine* or canine* or pet or pets or dolphin* or guinea pig* or rat or rats or rabbit* or chinchilla* or hamster*) adj1 (interaction* or activit* or therap* or intervention*)).tw,kw.
7 or/1-6
8 exp student/ or exp school/ or education/ or academic achievement/ or curriculum/ or learning environment/ or vocational education/
9 (school* or student* or classroom* or universit* or college* or education* or curricul* or campus* or diploma* or baccalaureate or higher stud* or graduat* or undergraduat* or facult* or learning environment or academic*).tw,kw.
10 8 or 9
11 7 and 10
12 limit 11 to conference abstracts
13 11 not 12

**Database: ERIC** 1965 to March 2020
Date searched: 10.06.2020
Number of hits: 145
Search Strategy:
1 ((dog* or cat or cats or horse* or equine* or canine* or dolphin* or guinea pig* or rat or rats or rabbit* or chinchilla* or hamster*) adj1 (assisted or facilitated or visitation*)).mp.
2 ((animal* or dog* or cat or cats or horse* or equine* or canine* or pet or pets or dolphin* or guinea pig* or rat or rats or rabbit* or chinchilla* or hamster*) adj1 therap*).mp.
3 (animal assisted or animal facilitated or animal visitation* or pet-facilitated or pet assisted or pet visitation* or hippotherapy).mp.
4 (service adj (animal* or dog* or cat or cats or horse* or equine* or canine* or pet or pets or dolphin* or guinea pig* or rat or rats or rabbit* or chinchilla* or hamster*)).mp.
5 ((human or people) adj1 (animal* or dog* or cat or cats or horse* or equine* or canine* or pet or pets or dolphin* or guinea pig* or rat or rats or rabbit* or chinchilla* or hamster*) adj1 (interaction* or activit* or therap* or intervention*)).mp.
6 or/1-5
7 (school* or student* or classroom* or universit* or college* or education* or curricul* or campus* or diploma* or baccalaureate or higher stud* or graduat* or undergraduat* or facult* or learning environment or academic*).mp.
8 schools/ or exp colleges/ or exp professional education/
9 7 or 8
10 6 and 9

**Database: APA PsycInfo** 1806 to June Week 1 2020
Date searched: 10.06.2020
Number of hits: 390
Search Strategy:
1 ((dog* or cat or cats or horse* or equine* or canine* or dolphin* or guinea pig* or rat or rats or rabbit* or chinchilla* or hamster*) adj1 (assisted or facilitated or visitation*)).mp.
2 ((animal* or dog* or cat or cats or horse* or equine* or canine* or pet or pets or dolphin* or guinea pig* or rat or rats or rabbit* or chinchilla* or hamster*) adj1 therap*).mp.
3 (animal assisted or animal facilitated or animal visitation* or pet-facilitated or pet assisted or pet visitation* or hippotherapy).mp.
4 (service adj (animal* or dog* or cat or cats or horse* or equine* or canine* or pet or pets or dolphin* or guinea pig* or rat or rats or rabbit* or chinchilla* or hamster*)).mp.
5 ((human or people) adj1 (animal* or dog* or cat or cats or horse* or equine* or canine* or pet or pets or dolphin* or guinea pig* or rat or rats or rabbit* or chinchilla* or hamster*) adj1 (interaction* or activit* or therap* or intervention*)).mp.
6 or/1-5
7 (school* or student* or classroom* or universit* or college* or education* or curricul* or campus* or diploma* or baccalaureate or higher stud* or graduat* or undergraduat* or facult* or learning environment or academic*).mp.
8 6 and 7
9 limit 8 to "0200 book"
10 8 not 9

**Database: CINAHL (Ebsco)**
Date searched: 10.06.2020
Number of hits: 447
Search Strategy:

S1 (MH "Pet Therapy") OR (MH "Equine-Assisted Therapy") OR (MH "Service Animals")

S2 TI ( ((dog* OR cat OR cats OR horse* OR equine* OR canine* OR dolphin* OR "guinea pig*" OR rat OR rats OR rabbit* OR chinchilla* OR hamster*) N0 (assisted OR facilitated OR visitation*)) ) OR AB ( ((dog* OR cat OR cats OR horse* OR equine* OR canine* OR dolphin* OR "guinea pig*" OR rat OR rats OR rabbit* OR chinchilla* OR hamster*) N0 (assisted OR facilitated OR visitation*)) )

S3 TI ( ((animal* OR dog* OR cat OR cats OR horse* OR equine* OR canine* OR pet OR pets OR dolphin* OR "guinea pig*" OR rat OR rats OR rabbit* OR chinchilla* OR hamster*) N0 therap*) ) OR AB ( ((animal* OR dog* OR cat OR cats OR horse* OR equine* OR canine* OR pet OR pets OR dolphin* OR "guinea pig*" OR rat OR rats OR rabbit* OR chinchilla* OR hamster*) N0 therap*) )

S4 TI ( (animal assisted OR animal facilitated OR animal visitation* OR pet-facilitated OR pet assisted OR pet visitation* OR hippotherapy) ) OR AB ( (animal assisted OR animal facilitated OR animal visitation* OR pet-facilitated OR pet assisted OR pet visitation* OR hippotherapy) )

S5 TI ( (service N0 (animal* OR dog* OR cat OR cats OR horse* OR equine* OR canine* OR pet OR pets OR dolphin* OR "guinea pig*" OR rat OR rats OR rabbit* OR chinchilla* OR hamster*)) ) OR AB ( (service N0 (animal* OR dog* OR cat OR cats OR horse* OR equine* OR canine* OR pet OR pets OR dolphin* OR "guinea pig*" OR rat OR rats OR rabbit* OR chinchilla* OR hamster*)) )

S6 TI ( ((human OR people) N0 (animal* OR dog* OR cat OR cats OR horse* OR equine* OR canine* OR pet OR pets OR dolphin* OR "guinea pig*" OR rat OR rats OR rabbit* OR chinchilla* OR hamster*) N0 (interaction* OR activit* OR therap* OR intervention*)) ) OR AB ( ((human OR people) N0 (animal* OR dog* OR cat OR cats OR horse* OR equine* OR canine* OR pet OR pets OR dolphin* OR "guinea pig*" OR rat OR rats OR rabbit* OR chinchilla* OR hamster*) N0 (interaction* OR activit* OR therap* OR intervention*)) )

S7 S1 OR S2 OR S3 OR S4 OR S5 OR S6

S8 (MH "Students") OR (MH "Student Dropouts") OR (MH "Students, College") OR (MH "Students, Disabled") OR (MH "Students, Foreign") OR (MH "Students, Health Occupations+") OR (MH "Education") OR (MH "Curriculum") OR (MH "Education, Health Sciences+") OR (MH "Faculty") OR (MH "Learning Environment+") OR (MH "Schools+") OR (MH "Student Assistance Programs") OR (MH "Student Experiences")

S9 TI ( (school* OR student* OR classroom* OR universit* OR college* OR education* OR curricul* OR campus* OR diploma* OR baccalaureate OR “higher stud*” OR graduat* OR undergraduat* OR facult* OR “learning environment” OR academic*) ) OR AB ( (school* OR student* OR classroom* OR universit* OR college* OR education* OR curricul* OR campus* OR diploma* OR baccalaureate OR “higher stud*” OR graduat* OR undergraduat* OR facult* OR “learning environment” OR academic*) )

S10 S8 OR S9

S11 S7 AND S10

**Database: Scopus (Elsevier)**
Date searched: 10.06.2020
Number of hits: 835
Search Strategy:
((TITLE-ABS-KEY(((dog* OR cat OR cats OR horse* OR equine* OR canine* OR dolphin* OR "guinea pig*" OR rat OR rats OR rabbit* OR chinchilla* OR hamster*) W/0 (assisted OR facilitated OR visitation*))) OR TITLE-ABS-KEY(((animal* OR dog* OR cat OR cats OR horse* OR equine* OR canine* OR pet OR pets OR dolphin* OR "guinea pig*" OR rat OR rats OR rabbit* OR chinchilla* OR hamster*) W/0 therap*)) OR TITLE-ABS-KEY(("animal assisted" OR "animal facilitated" OR "animal visitation*" OR "pet-facilitated" OR "pet assisted" OR "pet visitation*" OR hippotherapy)) OR TITLE-ABS-KEY((service W/0 (animal* OR dog* OR cat OR cats OR horse* OR equine* OR canine* OR pet OR pets OR dolphin* OR "guinea pig*" OR rat OR rats OR rabbit* OR chinchilla* OR hamster*))) OR TITLE-ABS-KEY((service PRE/0 (animal* OR dog* OR cat OR cats OR horse* OR equine* OR canine* OR pet OR pets OR dolphin* OR "guinea pig*" OR rat OR rats OR rabbit* OR chinchilla* OR hamster*))))) AND (TITLE-ABS-KEY((school* OR student* OR classroom* OR universit* OR college* OR education* OR curricul* OR campus* OR diploma* OR baccalaureate OR "higher stud*" OR graduat* OR undergraduat* OR facult* OR "learning environment" OR academic*)))

**Database: Web of Science**
Indexes=SCI-EXPANDED, SSCI, A&HCI, CPCI-S, CPCI-SSH, ESCI Timespan=1900-2020
Date searched: 10.06.2020
Number of hits: 509
Search Strategy:
TS=((dog* OR cat OR cats OR horse* OR equine* OR canine* OR dolphin* OR OR "guinea pig*" OR rat OR rats OR rabbit* OR chinchilla* OR hamster*) NEAR/0 (assisted OR facilitated OR visitation*) ) OR
TS=((assisted OR facilitated OR visitation*) NEAR/0 (dog* OR cat OR cats OR horse* OR equine* OR canine* OR dolphin* OR "guinea pig*" OR rat OR rats OR rabbit* OR chinchilla* OR hamster*) ) OR
TS=((animal* OR dog* OR cat OR cats OR horse* OR equine* OR canine* OR pet OR pets OR dolphin* OR "guinea pig*" OR rat OR rats OR rabbit* OR chinchilla* OR hamster*) NEAR/0 therap*) OR
TS=(therapy* NEAR/0 (animal* OR dog* OR cat OR cats OR horse* OR equine* OR canine* OR pet OR pets OR dolphin* OR "guinea pig*" OR rat OR rats OR rabbit* OR chinchilla* OR hamster*) ) OR
TS=("animal assisted" OR "animal facilitated" OR "animal visitation*" OR "pet-facilitated" OR "pet assisted" OR "pet visitation*" OR hippotherapy) OR
TS=(service NEAR/0 (animal* OR dog* OR cat OR cats OR horse* OR equine* OR canine* OR pet OR pets OR dolphin* OR "guinea pig*" OR rat OR rats OR rabbit* OR chinchilla* OR hamster*) ) OR
TS=((human OR people) NEAR/0 (animal* OR dog* OR cat OR cats OR horse* OR equine* OR canine* OR pet OR pets OR dolphin* OR "guinea pig*" OR rat OR rats OR rabbit* OR chinchilla* OR hamster*) NEAR/0 (interaction* OR activit* OR therap* OR intervention*) ) OR
TS=((animal* OR dog* OR cat OR cats OR horse* OR equine* OR canine* OR pet OR pets OR dolphin* OR "guinea pig*" OR rat OR rats OR rabbit* OR chinchilla* OR hamster*) NEAR/0 (human OR people) NEAR/0 (interaction* OR activit* OR therap* OR intervention*) )
AND
TS=(school* OR student* OR classroom* OR universit* OR college* OR education* OR curricul* OR campus* OR diploma* OR baccalaureate OR "higher stud*" OR graduat* OR undergraduat* OR facult* OR "learning environment" OR academic*)

**Database: Open Grey**
Date searched: 17.06.2020
Number of hits: 212 (with internal dublets)
Search Strategies:
(school* OR student* OR classroom* OR universit* OR college* OR education* OR curricul* OR campus* OR diploma* OR baccalaureate OR higher stud* OR graduat* OR undergraduat* OR facult* OR learning environment OR academic*) AND ((dog* OR cat OR cats OR horse* OR equine* OR canine* OR dolphin* OR "guinea pig*" OR rat OR rats OR rabbit* OR chinchilla* OR hamster*) NEAR/1 (assisted OR facilitated OR visitation*))
– 4 matches

(school* OR student* OR classroom* OR universit* OR college* OR education* OR curricul* OR campus* OR diploma* OR baccalaureate OR higher stud* OR graduat* OR undergraduat* OR facult* OR “learning environment” OR academic*) AND (animal* OR dog* OR cat OR cats OR horse* OR equine* OR canine* OR pet OR pets OR dolphin* OR "guinea pig*" OR rat OR rats OR rabbit* OR chinchilla* OR hamster*) NEAR/1 therap*)
– 19 matches

(school* OR student* OR classroom* OR universit* OR college* OR education* OR curricul* OR campus* OR diploma* OR baccalaureate OR higher stud* OR graduat* OR undergraduat* OR facult* OR learning environment OR academic*) AND (“animal assisted” OR “animal facilitated” OR “animal visitation*” OR “pet-facilitated” OR “pet assisted” OR “pet visitation*” OR hippotherapy)
– 44 matches

(school* OR student* OR classroom* OR universit* OR college* OR education* OR curricul* OR campus* OR diploma* OR baccalaureate OR higher stud* OR graduat* OR undergraduat* OR facult* OR “learning environment” OR academic*) AND (service NEAR/1 (animal* OR dog* OR cat OR cats OR horse* OR equine* OR canine* OR pet OR pets OR dolphin* OR "guinea pig*" OR rat OR rats OR rabbit* OR chinchilla* OR hamster*))
– 0 matches

(school* OR student* OR classroom* OR college* OR education* OR curricul* OR campus* OR diploma* OR baccalaureate OR higher stud* OR graduat* OR undergraduat* OR facult* OR “learning environment” OR academic*) AND (“human animal” OR “human dog” OR “human cat” OR “human horse” OR “human equine” OR “human canine” OR “human pet” OR “human dolphin” OR “human guinea pig*" OR “human rat” OR “human rats” OR “human rabbit*” OR “human chinchilla*” OR “human hamster*”) NEAR/1 (interaction* OR activit* OR therap* OR intervention*)
– 145 matches

**Database: WALTHAM Scientific Publications (accessible via https://www.waltham.com/resources/waltham-publications/)**
Date searched: 17.06.2020
Number of hits: 31
Search Strategy:
All publications in the sub-section “Human animal interaction” were included.

**Database: HABRI Central: Resources for the Study of the Human-Animal Bond (accessible via https://habricentral.org/resources/)**
Date searched: 19.06.2020
Number of hits: 410
Search Strategy:
After several trial searches to see which search terms yielded the most relevant results, the website was searched using the search term “students”. The results were filtered to show only resources in the form of journal articles, reports and theses.

**Database: Animals and Society Institute: Resources for Scholars and Researchers (accessible via https://www.animalsandsociety.org/human-animal-studies/faculty/)**
Date searched: 20.06.2020
Number of hits: 95
Search Strategy:
On the website listed above, the sub-section “Society and Animals articles by topic” (https://www.animalsandsociety.org/human-animal-studies/society-and-animals-journal/society-animals-articles-by-topic/) was searched for relevant literature. On the “Society and Animals articles by topic” website, all articles listed under the topics “Animal-assisted activities” (20 results), “Benefits of animals to humans” (17 results), and “Dogs” (58 results) were included.

**Explanations of the search history:**
* truncation
*Unlimited right-hand truncation* searches for variations on a word that are formed with different suffixes. For example, a search for *gene** finds occurrences of *gene*, *genes*, *genetics*, and *generation*.
Words with a **slash at the end (/)** is MeSH – medical subject headings (controlled vocabulary).
ADJn is a positional operator that lets you retrieve records that contain your terms (in any order) within a specified number (n) of words of each other. To apply adjacency, separate your search terms with the ADJ operator and a number from 1 to 99. The ADJ operators finds two terms next to each other in the specified order. The ADJ1 operators finds two terms next to each other in any order. The ADJ2 operator finds terms in any order and with one word (or none) between them. The ADJ3 operator finds terms in any order with two words (or fewer) between them. The ADJ4 operator finds terms in any order and with three words (or fewer) between them, and so on.
**Adj** between words defines how many words (any words) there can be between two words. Adj means **o, zero** words between the two words (for example: dog assisted.tw,kf.   = dog adj assisted.tw,kf.). **Adj1** means no words in between two other words, but the order of the words is interchangeable.
**tw,kf** = title field, abstract field or authors keyword field.
