## Supplementary material for "Animals in higher education settings: Do animal-assisted interventions improve mental and cognitive health outcomes of students? A systematic review and meta-analysis": S2_Table

| **Database searched** | **Number of retrieved references** |
|---|---|
| Medline (Ovid): | 307 |
| Embase (Ovid): | 378 |
| ERIC (Ovid) | 145 |
| PsycInfo (Ovid): | 390 |
| CINAHL (Ebsco): | 447 |
| Scopus | 835 |
| Web of Science (WoS) | 509 |
| OpenGrey (with duplicates) | 212 |
| WALTHAM Scientific Publications | 31 |
| | 95 |
| **Number of references before deduplication:** | **3759** |
| **Number of references after deduplication:** | **2431** |
