## Supplementary material for "Animals in higher education settings: Do animal-assisted interventions improve mental and cognitive health outcomes of students? A systematic review and meta-analysis": S3_File

- Study title
- Study authors and year
- Study design
  - Study design (RCT or crossover RCT)
  - Type of publication (peer-reviewed journal article or undergraduate/postgraduate/doctoral dissertation)
- Participants
  - Gender
  - Age
  - Number of participants
  - Were sample size calculations conducted?
  - Study location
  - If applicable: pre-existing health condition
  - Were participants pet owners?
  - University degree (subject)
  - University level (Bachelor, Master, PhD)
  - Recruitment process
  - Eligibility criteria for participants
- Intervention
  - Randomization process into intervention and control groups
  - Differences between intervention and control group at baseline
  - Purpose of intervention (physical therapy/stress prevention/anxiety reduction etc.)
  - Was intervention an AAI program integrated in a pre-existing university program?
  - Number of participants in intervention condition
  - Animals in intervention condition
    - Type of animal
    - Breed of animal
    - Training level of animal
    - Number of animals present in intervention condition
  - If applicable: Person administering intervention (researcher, animal handler etc.)
  - If applicable: number of handlers present in intervention condition
  - Animal-to-participant ratio
  - Description of intervention condition
    - Type of activity (presence of animal, active interaction with animal etc.)
    - If applicable: measures used to induce stress among participants (test, interview etc.)
    - Setting of intervention condition
    - Format of intervention condition (groups, individual)
    - Timing of intervention (when in the semester did it take place? Beginning, middle, end, before exams, after exams?)
    - Duration of intervention condition
    - Number of intervention sessions per participant
    - Frequency of individual intervention sessions per participant
    - Duration of individual intervention sessions
- If applicable: Control
  - Number of control conditions
  - Number of participants in control condition(s)
  - Description of control condition(s)
    - Type of activity

        - Setting of control condition
        - Format of control condition (groups, individual)
        - Timing of control (when in the semester did it take place? Beginning, middle, end, before exams, after exams?)
        - Duration of control condition
        - Number of control sessions per participant
        - Frequency of individual control session per participant
        - Duration of individual control session
- Outcomes
    - Quantitative outcomes
        - Outcomes assessed
        - Measures used to assess outcomes
        - Timing of outcome assessments (baseline, immediate at the start of/during intervention, immediately following intervention, long-term)
        - Statistical test used
        - Results from quantitative outcome assessment
        - If available: effect size (Hedge's g) and corresponding SE
        - P-value
        - If applicable: other relevant outcomes from sub-group analyses
    - Qualitative outcomes
        - Outcomes assessed
        - Measures used to assess outcomes
        - Timing of outcome assessments (baseline, immediate at the start of/during intervention, immediately following intervention, long-term)
        - Results from qualitative outcome assessment
