## Supplementary material for "Animals in higher education settings: Do animal-assisted interventions improve mental and cognitive health outcomes of students? A systematic review and meta-analysis": S4_Table

| Reference | Outcomes included in meta-analysis | Input used („esc“ function used) | Hedges‘ g and SE |
|---|---|---|---|
| Banks et al. (2018) | Chronic self-perceived stress<br><br>Acute anxiety<br><br>Positive affect<br><br>Negative affect | means and sds (esc_mean_sd) | Chronic self-perceived stress: g= -0.411, SE= 0.27<br><br>Acute anxiety: g= -0.52, SE= 0.272<br><br>Positive affect: g= -0.104, SE= 0.268<br><br>Negative affect: g= -0.011, SE= 0.267 |
| Binfet et al. (2017) | Chronic self-perceived stress | means and sds (esc_mean_sd) | g= -0.326, SE= 0.162 |
| Crossman et al. (2015) | Acute anxiety<br><br>Positive affect<br><br>Negative affect | means and sds (esc_mean_sd) | Acute anxiety: g= -1.245, SE= 0.331<br><br>Negative affect: g= -1.391, SE=0.337<br><br>Positive affect: g= 0.564, SE= 0.308 |
| Crump et al. (2015) - Study I | Systolic blood pressure | f-test (esc_f) | Systolic blood pressure: g= 0.814, SE= 0.402 |
| McDonald et al. (2017) | Systolic blood pressure | means and sds (esc_mean_sd) | g= -2.501, SE= 0.389 |
| Shearer et al. (2015) | Acute anxiety<br><br>Negative affect | means and sds at time point 4 (esc_mean_sd) | Acute anxiety: g= -0.811, SE= 0.328<br><br>Negative affect: g= -0.612, SE= 0.322 |
| Ward-Griffin et al. (2018) - RCT | Positive affect<br><br>Negative affect<br><br>Chronic self-perceived stress | means and sds (esc_mean_sd) | Positive affect: g= -0.088, SE= 0.128<br><br>Negative affect: g= -0.081, SE= 0.127<br><br>Chronic self-perceived stress: g= -0.128, SE= 0.128 |
| Wilson (1987) | Systolic blood pressure<br><br>Acute anxiety | means and sds (esc_mean_sd) (intervention vs. reading quietly) | Systolic blood pressure: g= 0.21, SE= 0.21<br><br>Acute anxiety: g= 0.0413, SE= 0.209 |
