## Supplementary material for "Animals in higher education settings: Do animal-assisted interventions improve mental and cognitive health outcomes of students? A systematic review and meta-analysis": S5_Table

| Reference | Gender | Location | Type of intervention condition | Type of control condition | Stressor?[a] | Format[b] | Frequency of sessions[c] | Duration of sessions (mins) |
|---|---|---|---|---|---|---|---|---|
| Banks et al. (2018) (113) | Mostly female | USA | Active | No treatment | Before exams | Group | 1 | 10 |
| Barker et al. (2016) (114) | Mostly female | USA | Active | No treatment | Before exams | Group | 1 | 15 |
| Barker et al. (2017) (115) | Mostly female | USA | Active | No treatment | Before exams | Group | 1 | 15 |
| Binfet et al. (2017) (126) | Mostly female | Canada | Active | No treatment | No | Group | 1 | 20 |
| Caparelli et al. (2020) (48) | Mostly female | USA | Passive | No treatment | No | Individual | 1 | 6 |
| Charnetski et al. (2004) (110) | n/s[4] | USA | Active | (1) animal, (2) no treatment | No | Individual | 1 | 18 |
| Crossman et al. (2015) (116) | Half male, half female | USA | Active | (1) animal, (2) no treatment | No | Individual | 1 | 7 |
| Crump et al. (2015) - Study I (102) | All female | USA | Active | No treatment | Before exams | Group | 1 | 15 |
| Crump et al. (2015) - Study II (102) | All female | USA | Active | No treatment | Before exams | Group | 1 | 30 |
| Fiocco & Hunse (2017) (105) | Mostly female | Canada | Active | No treatment | Yes | Individual | 1 | 10 |
| Gebhart et al. (2019) (127) | Mostly female | Austria | Active | (1 and 2) other, (3) no treatment | Before exams | Group | >1 | 45 |
| Gee et al. (2014) (106) | Mostly female | USA | Passive | (1) human, (2) animal | Yes | Individual | 1 | 3 |
| Gee et al. (2015) (117) | Mostly female | USA | Passive | (1 and 2) human, (3) no treatment | Yes | Individual | 1 | 3 |
| Gee et al. (2019) - Experiment 1 (104) | Mostly female | USA | Active | (1) animal, (2) no treatment | Yes | Individual | 1 | 5 |
| Gee et al. (2019) - Experiment 2 (104) | Mostly female | USA | Active | (1) animal, (2) no treatment | Yes | Individual | 1 | 5 |
| Gonzalez-Ramirez et al. (2016) (130) | Half male, half female | Mexico | Active | No treatment | Yes | Group | n/s | n/s |
| Grajfoner et al. (2017) (128) | Mostly female | Scotland | Active | (1) animal, (2) human | No | Group | 1 | 20 |

| | | | | | | | | |
|---|---|---|---|---|---|---|---|---|
| Hall (2018) (118) | Mostly female | USA | Active | No treatment | No | Group or individual | n/s | n/s |
| Hunt & Chizkov (2014) (109) | Mostly female | USA | Passive | (1 and 2) no treatment | Yes | Individual | >1 | 20 |
| Kobayashi et al. (2017) (129) | Mostly female | Japan | Active | Animal | No | Individual | 1 | 1 |
| McDonald et al. (2017) (119) | n/s | USA | Active | No treatment | Before exams | Group | 1 | 15 |
| Pendry & Vandagriff (2019) (120) | Mostly female | USA | Active | (1) animal, (2 and 3) no treatment | Before exams | Dogs: group, cats: individual | 1 | 10 |
| Pendry et al. (2018) (121) | Mostly female | USA | Active | (1) animal, (2) no treatment | Before exams | Dogs: group, cats: individual | 1 | 10 |
| Pendry et al. (2019) (122) | Mostly female | USA | Active | (1 and 2) other | No | Group | >1 | 60 |
| Pendry et al. (2019, Clinical depression) (60) | Mostly female | USA | Active | (1) animal, (2) no treatment | Before exams | Dogs: group, cats: individual | 1 | 10 |
| Pendry et al. (2020) (36) | Mostly female | USA | Active | (1 and 2) other | No | Group | >1 | 60 |
| Polheber & Matchock (2014) (108) | Half male, half female | USA | Passive | (1) human, (2) no treatment | Yes | Individual | 1 | 53 |
| Shearer et al. (2015) (123) | Half male, half female | USA | Active | (1) other, (2) no treatment | No | Group | >1 | 60 |
| Stewart & Strickland (2013) (107) | Mostly female | USA | Passive | No treatment | Yes | Individual | 1 | 14 |
| Straatman et al. (1997) (111) | All male | Netherlands | Passive | No treatment | Yes | Individual | 1 | 36 |
| Trammell (2017) - Study 2 (103) | n/s | USA | Active | Animal | Before exams | Group | 1 | 15 |
| Trammell (2017) - Study 3 (103) | n/s | USA | Active | Animal | Before exams | Group | 1 | 15 |
| Trammell (2019) (124) | Mostly female | USA | Passive | No treatment | Yes | Group | >1 | n/s |
| Ward-Griffin et al. (2018) (125) | Mostly female | Canada | Active | No treatment | No | Group | 1 | 30 |
| Wilson (1987) (112) | Mostly female | USA | Active | (1) other, (2) no treatment | No | Individual | 1 | 10 |
