## Supplementary figures and images for "Animals in higher education settings: Do animal-assisted interventions improve mental and cognitive health outcomes of students? A systematic review and meta-analysis"

### S6_Figure

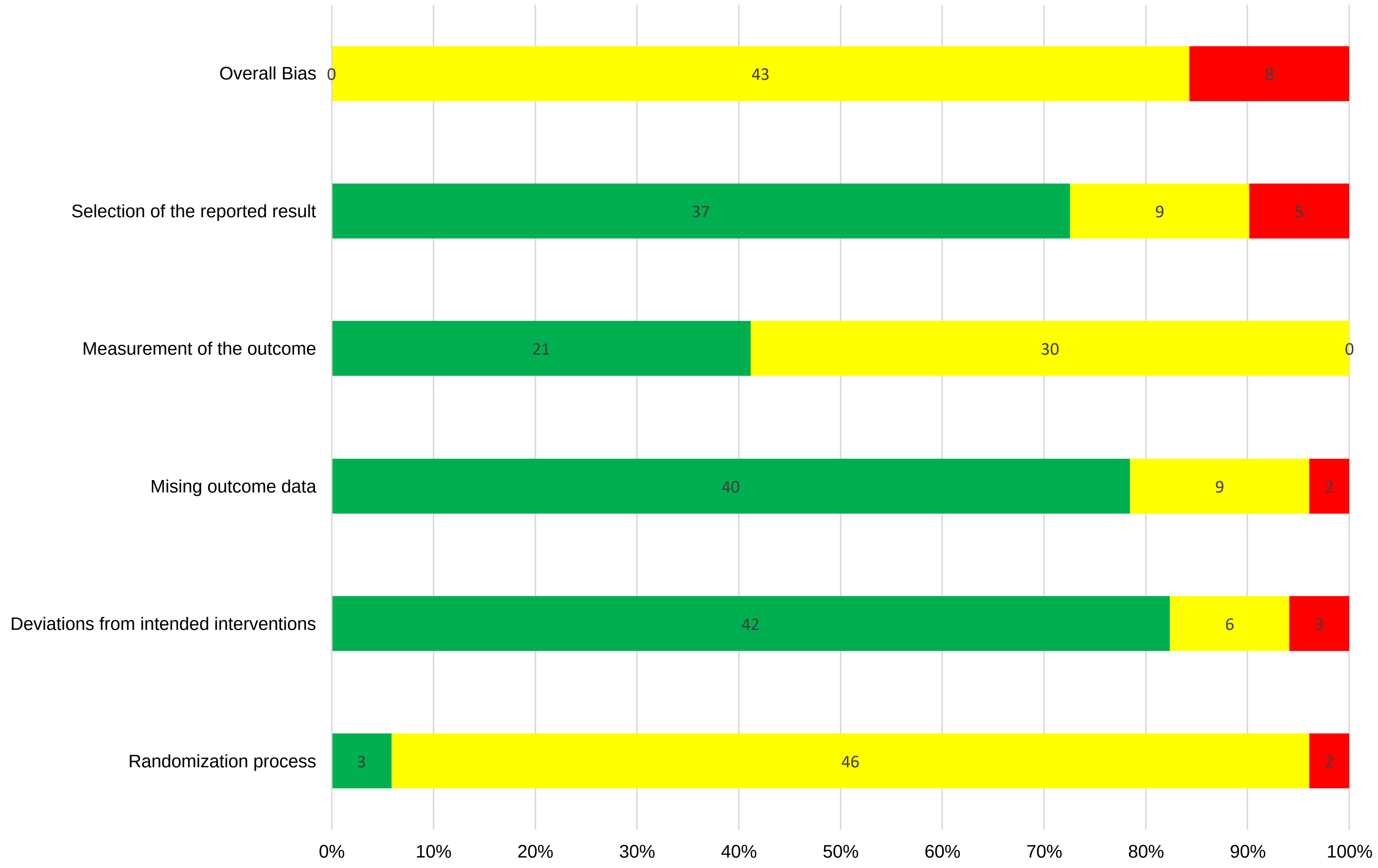

### S7_Figure

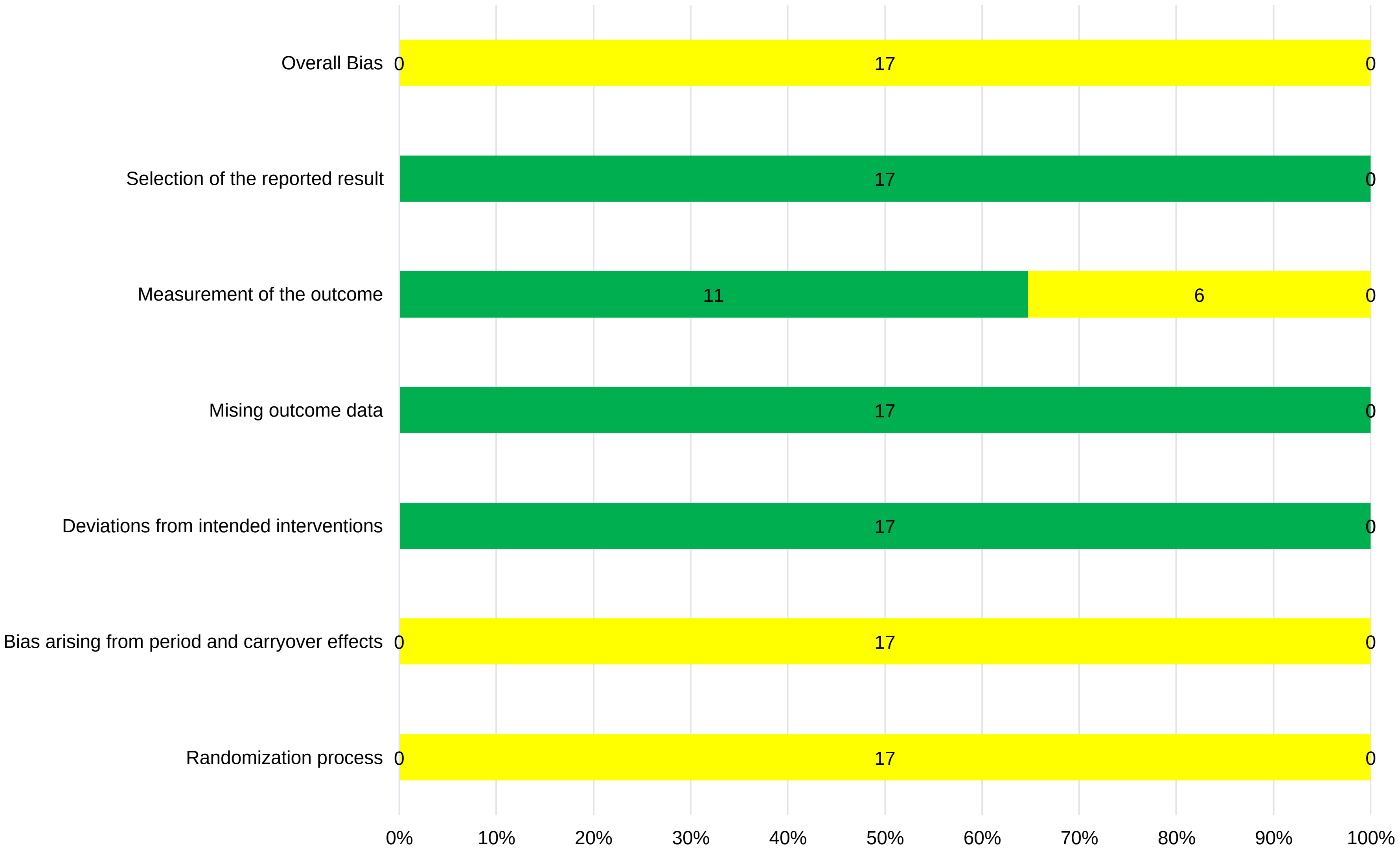

### S9_Figure

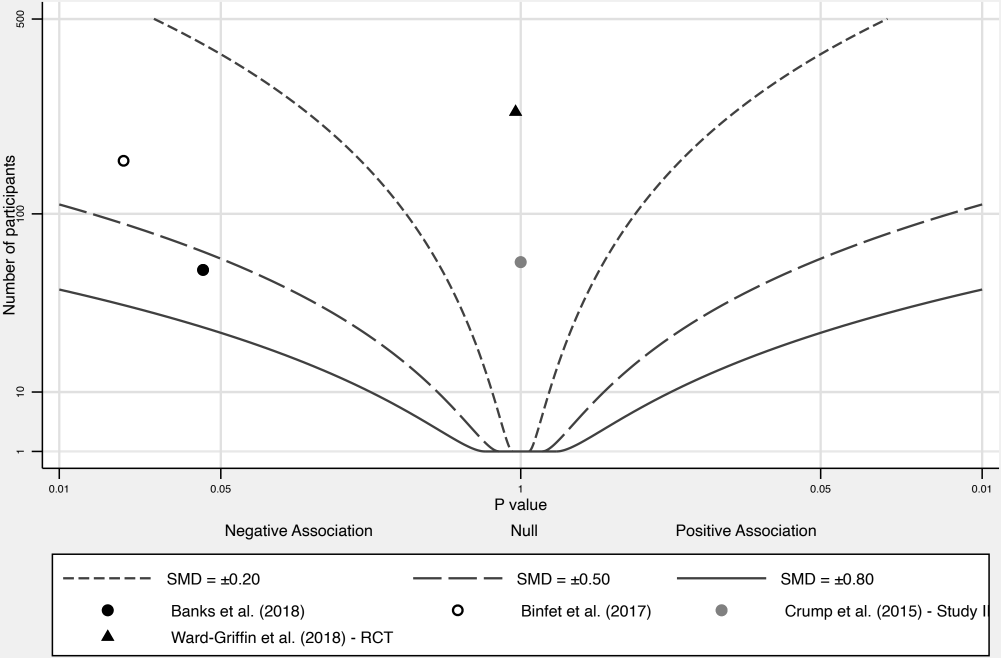


Effect contours drawn using a ratio of group sizes (r) of 1.00

Grouped by: Studyauthorsandyear

### S10_Figure

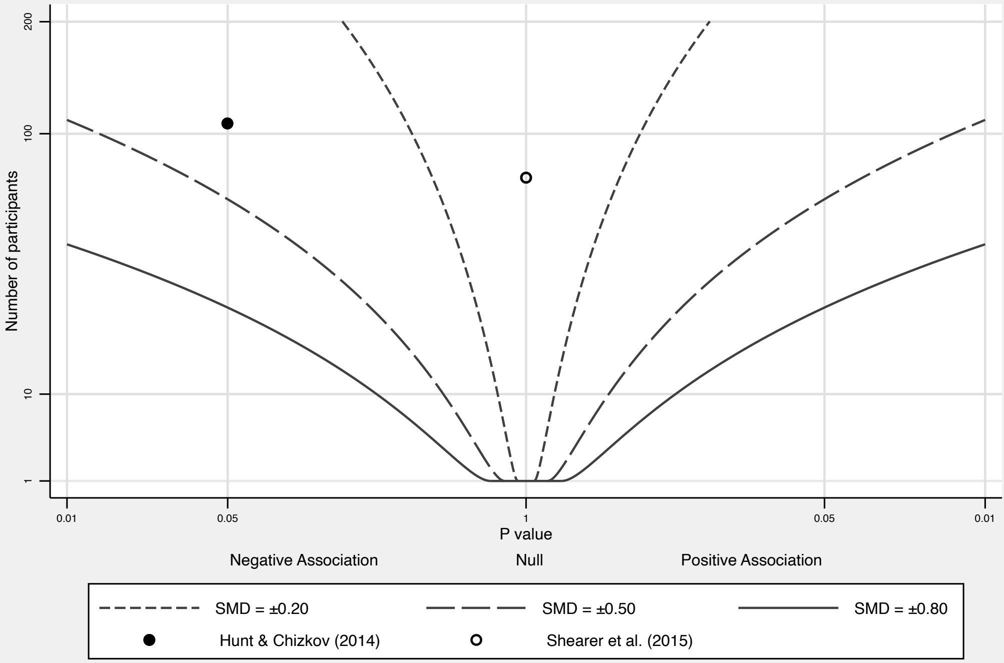


Effect contours drawn using a ratio of group sizes (r) of 1.00

Grouped by: Studyauthorsandyear

### S11_Figure

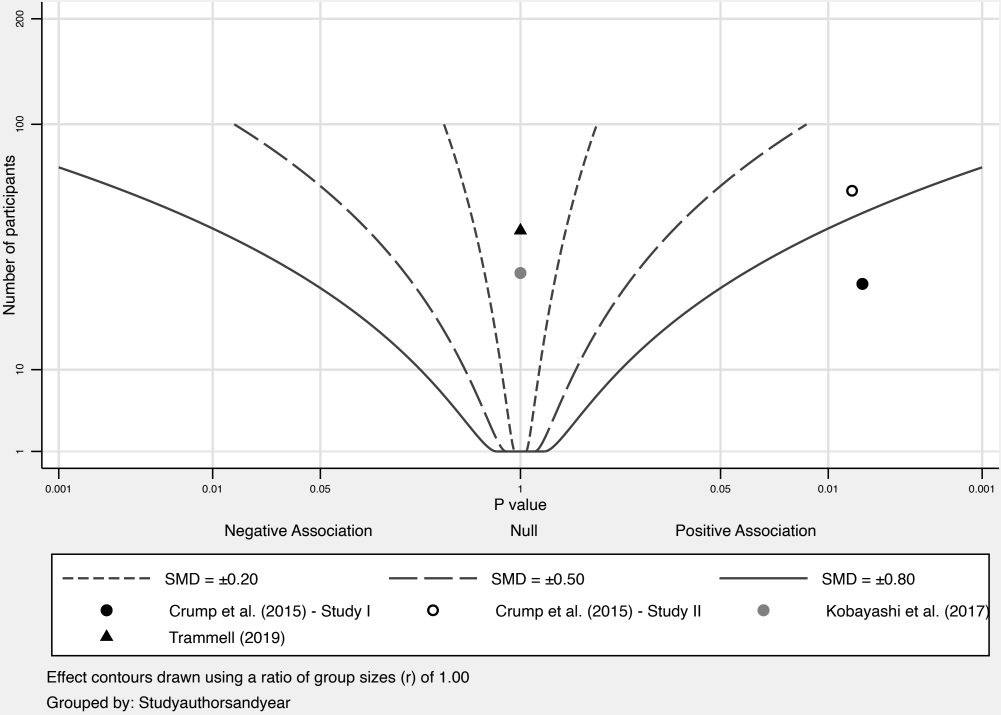

### S12_Figure

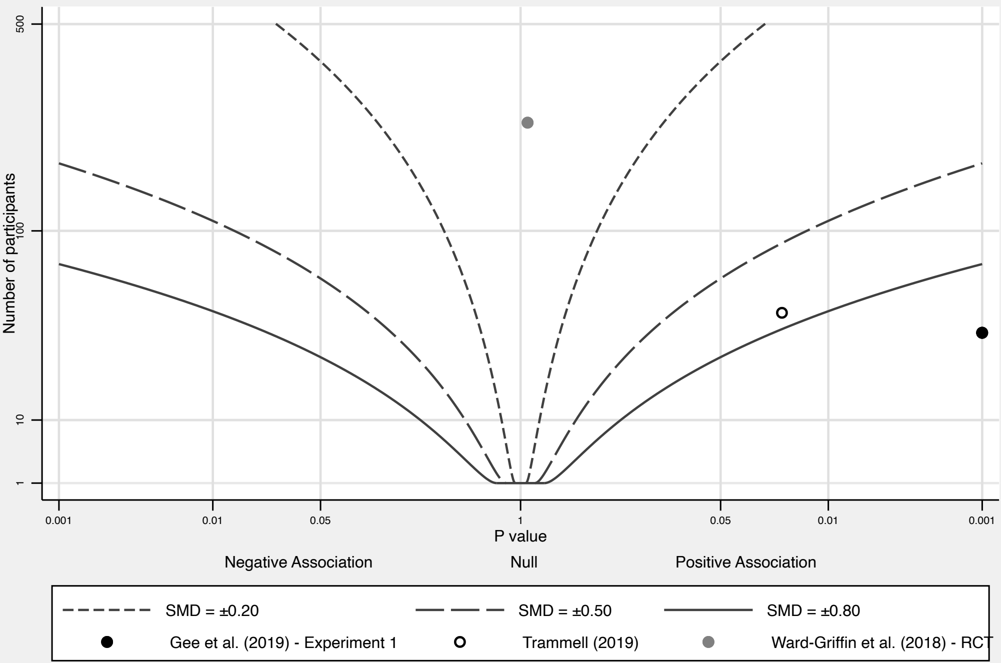


Effect contours drawn using a ratio of group sizes (r) of 1.00

Grouped by: Studyauthorsandyear

### S13_Figure

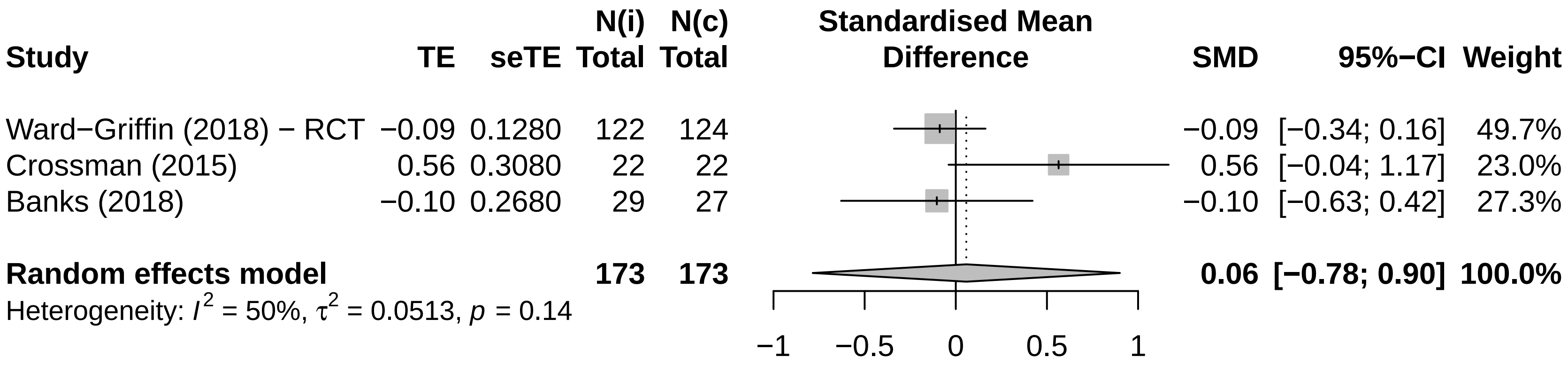

### S14_Figure

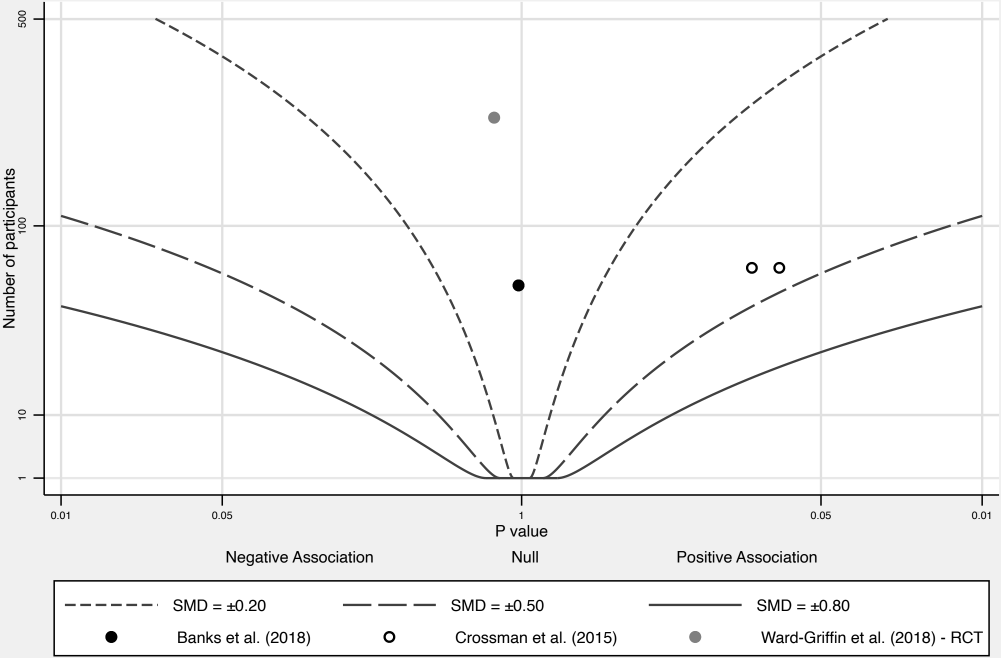


Effect contours drawn using a ratio of group sizes (r) of 1.00

Grouped by: Studyauthorsandyear

### S15_Figure

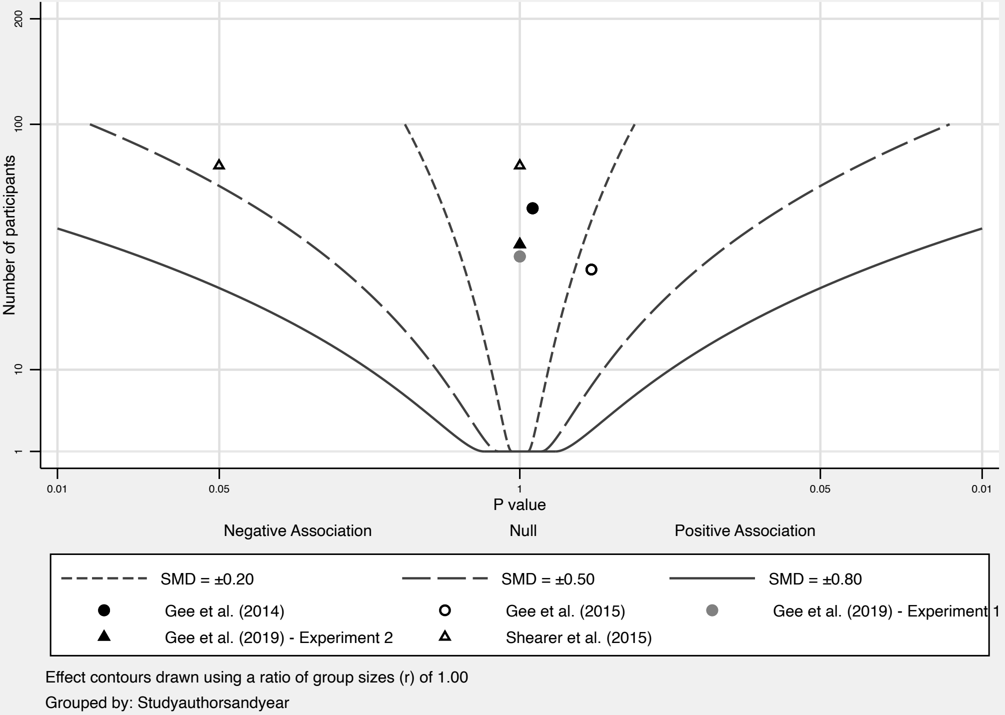

### S16_Figure

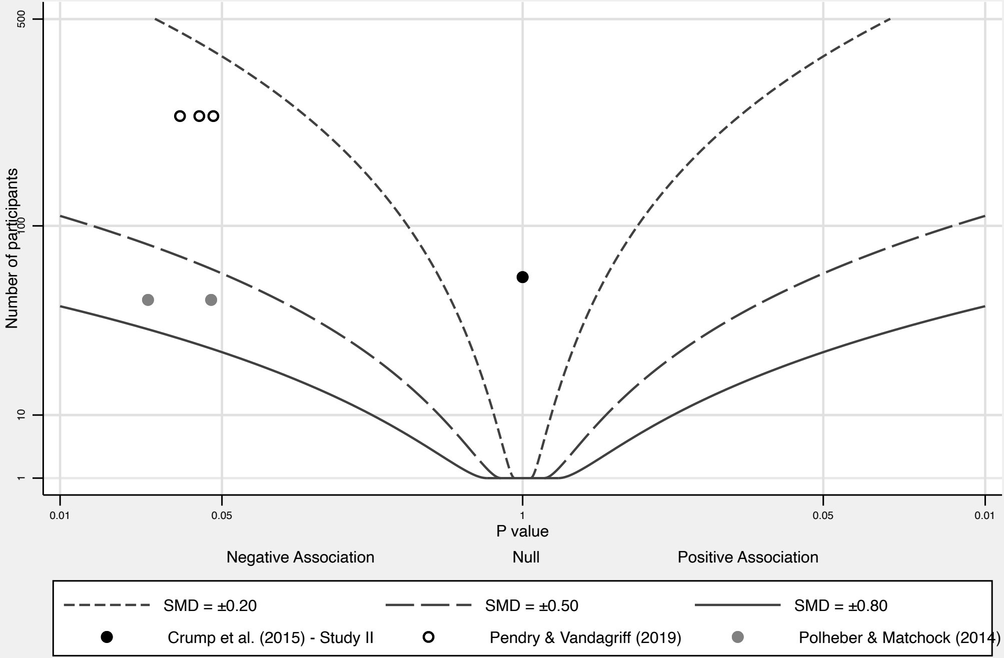


Effect contours drawn using a ratio of group sizes (r) of 1.00

Grouped by: Studyauthorsandyear

### S18_Figure

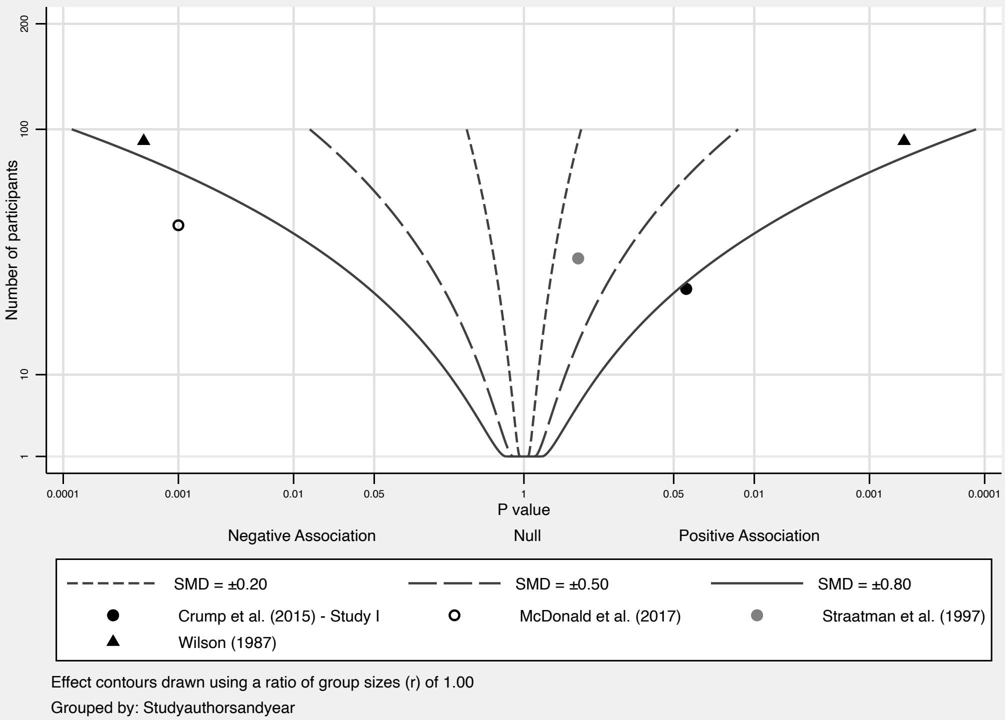

### S19_Figure

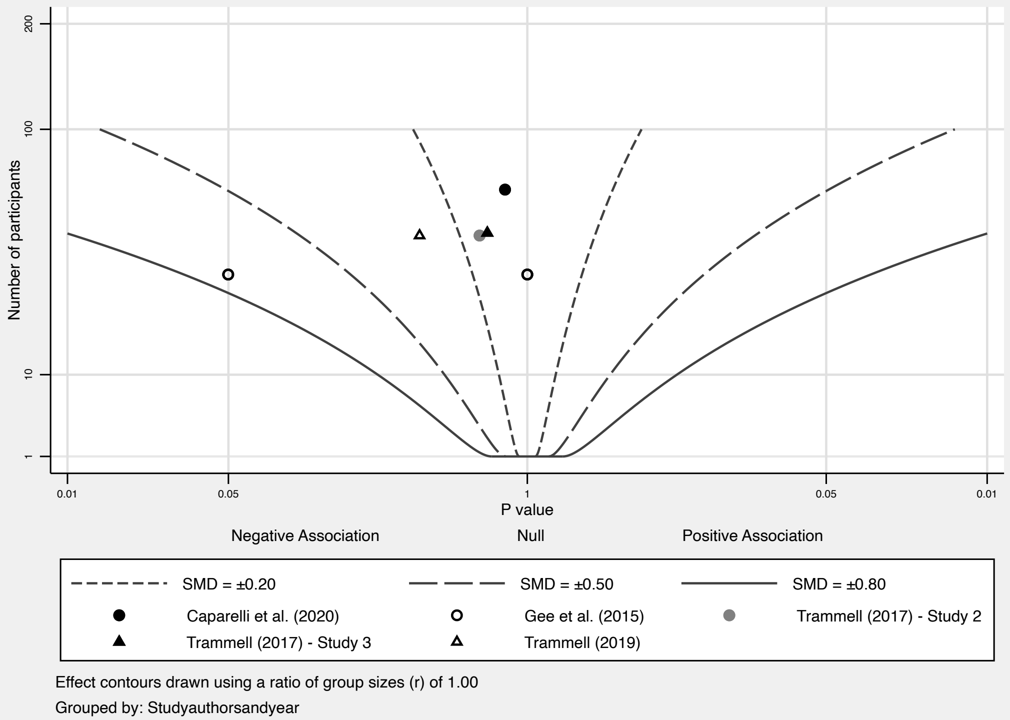

### S20_Figure

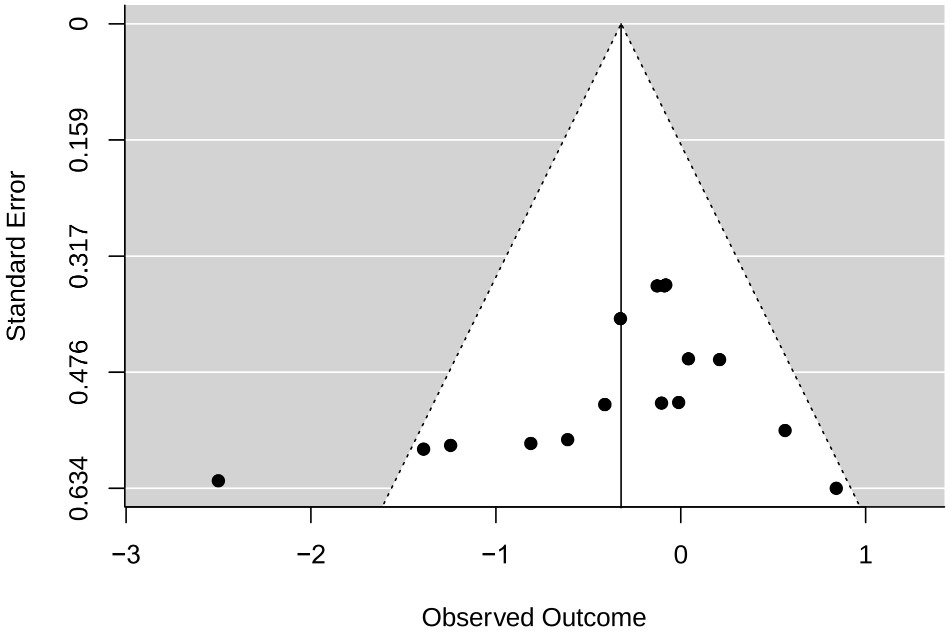
