## Supplementary material for "Animals in higher education settings: Do animal-assisted interventions improve mental and cognitive health outcomes of students? A systematic review and meta-analysis": S6_Table

| Reference | Outcome | Effect of adhering to intervention? | Randomization process | Deviations from intended interventions | Mising outcome data | Measurement of the outcome | Selection of the reported result | Overall Bias |
|---|---|---|---|---|---|---|---|---|
| **Banks et al. (2018)** | Chronic self-perceived stress | Effect of assignment to intervention | Some concerns | Low risk | Low risk | Some concerns | Low risk | Some concerns |
| Banks et al. (2018) | Positive and negative affect | Effect of assignment to intervention | Some concerns | Low risk | Low risk | Some concerns | Low risk | Some concerns |
| Banks et al. (2018) | Acute anxiety | Effect of assignment to intervention | Some concerns | Low risk | Low risk | Some concerns | Low risk | Some concerns |
| **Binfet et al. (2017)** | Chronic self-perceived stress | Effect of assignment to intervention | Some concerns | Low risk | Some concerns | Some concerns | Low risk | Some concerns |
| **Caparelli et al. (2020)** | Memory test | Effect of assignment to intervention | Some concerns | Some concerns | Low risk | Low risk | Some concerns | Some concerns |
| **Crossman et al. (2015)** | Acute anxiety | Effect of assignment to intervention | Some concerns | Low risk | Low risk | Some concerns | Low risk | Some concerns |
| Crossman et al. (2015) | Positive and negative affect | Effect of assignment to intervention | Some concerns | Low risk | Low risk | Some concerns | Low risk | Some concerns |
| **Crump et al. (2015) - Study II** | Acute self-perceived stress | Effect of assignment to intervention | Some concerns | Low risk | Low risk | Some concerns | Low risk | Some concerns |
| Crump et al. (2015) - Study II | Arousal | Effect of assignment to intervention | Some concerns | Low risk | Low risk | Some concerns | Low risk | Some concerns |
| Crump et al. (2015) - Study II | Chronic self-perceived stress | Effect of assignment to intervention | Some concerns | Low risk | Low risk | Some concerns | Low risk | Some concerns |
| Crump et al. (2015) - Study II | Salivary cortisol | Effect of assignment to intervention | Some concerns | Low risk | Low risk | Low risk | Low risk | Some concerns |
| **Fiocco & Hunse (2017)** | Positive and negative affect | Effect of assignment to intervention | Some concerns | Low risk | Low risk | Some concerns | Low risk | Some concerns |
| **Gebhart et al. (2018)** | Acute anxiety | Effect of assignment to intervention | Low risk | Low risk | Some concerns | Some concerns | Low risk | Some concerns |
| Gebhart et al. (2018) | Acute self-perceived stress | Effect of assignment to intervention | Low risk | Low risk | Some concerns | Some concerns | Low risk | Some concerns |
| Gebhart et al. (2018) | Salivary cortisol | Effect of assignment to intervention | Low risk | Low risk | Some concerns | Low risk | Low risk | Some concerns |
| **Gee et al. (2019) - Experiment 2** | Acute anxiety | Effect of assignment to intervention | Some concerns | Low risk | Low risk | Some concerns | Low risk | Some concerns |
| Gee et al. (2019) - Experiment 2 | HR | Effect of assignment to intervention | Some concerns | Low risk | Low risk | Low risk | Low risk | Some concerns |
| Gee et al. (2019) - Experiment 2 | HRV | Effect of assignment to intervention | Some concerns | Low risk | Low risk | Low risk | Low risk | Some concerns |
| **Grajfoner et al. (2017)** | Acute anxiety | Effect of assignment to intervention | Some concerns | Low risk | Low risk | Some concerns | Low risk | Some concerns |
| **Hall (2018)** | Acute anxiety | Effect of assignment to intervention | Some concerns | High risk | Some concerns | Some concerns | Low risk | High risk |
| **Hunt & Chizkov (2015)** | Chronic depression | Effect of assignment to intervention | Some concerns | Low risk | Low risk | Some concerns | High risk | High risk |
| Hunt & Chizkov (2015) | Acute anxiety | Effect of assignment to intervention | Some concerns | Low risk | Low risk | Some concerns | High risk | High risk |

| | | | | | | | | |
|---|---|---|---|---|---|---|---|---|
| Hunt & Chizkov (2015) | Positive and negative affect | Effect of assignment to intervention | Some concerns | Low risk | Low risk | Some concerns | High risk | High risk |
| **McDonald et al. (2017)** | BP | Effect of assignment to intervention | Some concerns | Some concerns | Low risk | Low risk | Some concerns | Some concerns |
| **Pendry & Vandagriff (2019)** | Salivary cortisol | Effect of assignment to intervention | Some concerns | Low risk | Low risk | Low risk | Low risk | Some concerns |
| **Pendry et al. (2018)** | Acute anxiety | Effect of assignment to intervention | High risk | High risk | High risk | Low risk | Low risk | High risk |
| Pendry et al. (2018) | Acute depression | Effect of assignment to intervention | High risk | High risk | High risk | Low risk | Low risk | High risk |
| **Pendry et al. (2019)** | Acute anxiety | Effect of assignment to intervention | Some concerns | Low risk | Low risk | Some concerns | Low risk | Some concerns |
| Pendry et al. (2019) | Acute depression | Effect of assignment to intervention | Some concerns | Low risk | Low risk | Some concerns | Low risk | Some concerns |
| **Polheber & Matchock (2014)** | HR | Effect of assignment to intervention | Some concerns | Low risk | Low risk | Low risk | Low risk | Some concerns |
| Polheber & Matchock (2014) | Salivary cortisol | Effect of assignment to intervention | Some concerns | Low risk | Low risk | Low risk | Low risk | Some concerns |
| Polheber & Matchock (2014) | Acute anxiety | Effect of assignment to intervention | Some concerns | Low risk | Low risk | Some concerns | Low risk | Some concerns |
| **Shearer et al. (2015)** | Acute anxiety | Effect of assignment to intervention | Some concerns | Some concerns | Some concerns | Some concerns | Some concerns | Some concerns |
| Shearer et al. (2015) | Negative affect | Effect of assignment to intervention | Some concerns | Some concerns | Some concerns | Some concerns | Some concerns | Some concerns |
| Shearer et al. (2015) | Chronic depression | Effect of assignment to intervention | Some concerns | Some concerns | Some concerns | Some concerns | High risk | High risk |
| Shearer et al. (2015) | HRV | Effect of assignment to intervention | Some concerns | Some concerns | Low risk | Low risk | Low risk | Some concerns |
| **Stewart & Strickland (2013)** | Acute anxiety | Effect of assignment to intervention | Some concerns | Low risk | Low risk | Low risk | Some concerns | Some concerns |
| **Straatman et al. (1997)** | HR | Effect of assignment to intervention | Some concerns | Low risk | Low risk | Low risk | Low risk | Some concerns |
| Straatman et al. (1997) | BP | Effect of assignment to intervention | Some concerns | Low risk | Low risk | Low risk | Low risk | Some concerns |
| Straatman et al. (1997) | Acute anxiety | Effect of assignment to intervention | Some concerns | Low risk | Low risk | Some concerns | High risk | High risk |
| **Trammell (2017) - Study 2** | Acute self-perceived stress | Effect of assignment to intervention | Some concerns | Low risk | Low risk | Some concerns | Low risk | Some concerns |
| Trammell (2017) - Study 2 | Memory test | Effect of assignment to intervention | Some concerns | Low risk | Low risk | Low risk | Low risk | Some concerns |
| **Trammell (2017) - Study 3** | Acute self-perceived stress | Effect of assignment to intervention | Some concerns | Low risk | Low risk | Some concerns | Low risk | Some concerns |
| Trammell (2017) - Study 3 | Memory test | Effect of assignment to intervention | Some concerns | Low risk | Low risk | Low risk | Low risk | Some concerns |
| **Trammell (2019)** | Memory test | Effect of assignment to intervention | Some concerns | Low risk | Low risk | Low risk | Some concerns | Some concerns |

| | | | | | | | | |
|---|---|---|---|---|---|---|---|---|
| Trammell (2019) | Happiness | Effect of assignment to intervention | Some concerns | Low risk | Low risk | Some concerns | Some concerns | Some concerns |
| Trammell (2019) | Acute self-perceived stress | Effect of assignment to intervention | Some concerns | Low risk | Low risk | Some concerns | Some concerns | Some concerns |
| Trammell (2019) | Arousal | Effect of assignment to intervention | Some concerns | Low risk | Low risk | Some concerns | Some concerns | Some concerns |
| **Ward-Griffin et al. (2018)** | Positive and negative affect | Effect of assignment to intervention | Some concerns | Low risk | Low risk | Low risk | Low risk | Some concerns |
| Ward-Griffin et al. (2018) | Chronic self-perceived stress | Effect of assignment to intervention | Some concerns | Low risk | Low risk | Low risk | Low risk | Some concerns |
| Ward-Griffin et al. (2018) | Happiness | Effect of assignment to intervention | Some concerns | Low risk | Some concerns | Low risk | Low risk | Some concerns |
