## Supplementary material for "Animals in higher education settings: Do animal-assisted interventions improve mental and cognitive health outcomes of students? A systematic review and meta-analysis": S7_Table

| Reference | Outcome | Effect of adhering to intervention? | Randomization process | Bias arising from period and carryover effects | Deviations from intended interventions | Mising outcome data | Measurement of the outcome | Selection of the reported result | Overall Bias |
|---|---|---|---|---|---|---|---|---|---|
| **Barker et al. (2016)** | Acute self-perceived stress | Effect of assignment to intervention | Some concerns | Some concerns | Low risk | Low risk | Some concerns | Low risk | Some concerns |
| **Crump et al. (2015) - Study I** | Acute self-perceived stress | Effect of assignment to intervention | Some concerns | Some concerns | Low risk | Low risk | Some concerns | Low risk | Some concerns |
| Crump et al. (2015) - Study I | Arousal | Effect of assignment to intervention | Some concerns | Some concerns | Low risk | Low risk | Some concerns | Low risk | Some concerns |
| Crump et al. (2015) - Study I | BP | Effect of assignment to intervention | Some concerns | Some concerns | Low risk | Low risk | Low risk | Low risk | Some concerns |
| Crump et al. (2015) - Study I | HR | Effect of assignment to intervention | Some concerns | Some concerns | Low risk | Low risk | Low risk | Low risk | Some concerns |
| **Gee et al. (2014)** | HR | Effect of assignment to intervention | Some concerns | Some concerns | Low risk | Low risk | Low risk | Low risk | Some concerns |
| Gee et al. (2014) | HRV | Effect of assignment to intervention | Some concerns | Some concerns | Low risk | Low risk | Low risk | Low risk | Some concerns |
| **Gee et al. (2015)** | HR | Effect of assignment to intervention | Some concerns | Some concerns | Low risk | Low risk | Low risk | Low risk | Some concerns |
| Gee et al. (2015) | HRV | Effect of assignment to intervention | Some concerns | Some concerns | Low risk | Low risk | Low risk | Low risk | Some concerns |
| Gee et al. (2015) | Memory test | Effect of assignment to intervention | Some concerns | Some concerns | Low risk | Low risk | Low risk | Low risk | Some concerns |
| **Gee et al. (2019) - Experiment 1** | Happiness | Effect of assignment to intervention | Some concerns | Some concerns | Low risk | Low risk | Some concerns | Low risk | Some concerns |
| Gee et al. (2019) - Experiment 1 | HR | Effect of assignment to intervention | Some concerns | Some concerns | Low risk | Low risk | Low risk | Low risk | Some concerns |
| Gee et al. (2019) - Experiment 1 | HRV | Effect of assignment to intervention | Some concerns | Some concerns | Low risk | Low risk | Low risk | Low risk | Some concerns |
| **Kobayashi et al. (2017)** | Arousal | Effect of assignment to intervention | Some concerns | Some concerns | Low risk | Low risk | Some concerns | Low risk | Some concerns |
| **Wilson (1987)** | BP | Effect of assignment to intervention | Some concerns | Some concerns | Low risk | Low risk | Low risk | Low risk | Some concerns |
| Wilson (1987) | HR | Effect of assignment to intervention | Some concerns | Some concerns | Low risk | Low risk | Low risk | Low risk | Some concerns |
| Wilson (1987) | Acute and chronic anxiety | Effect of assignment to intervention | Some concerns | Some concerns | Low risk | Low risk | Some concerns | Low risk | Some concerns |
