## Supplementary material for "Animals in higher education settings: Do animal-assisted interventions improve mental and cognitive health outcomes of students? A systematic review and meta-analysis": S8_Figure

| Study | TE | seTE | N(i) Total | N(c) Total | Standardised Mean Difference | SMD | 95%−CI | Weight |
|---|---|---|---|---|---|---|---|---|
| Ward−Griffin (2018) − RCT | −0.13 | 0.1280 | 122 | 124 | | −0.13 | [−0.38; 0.12] | 54.1% |
| Banks (2018) | −0.41 | 0.2700 | 29 | 27 | | −0.41 | [−0.94; 0.12] | 12.2% |
| Binfet (2017) | −0.33 | 0.1620 | 84 | 79 | | −0.33 | [−0.64; −0.01] | 33.8% |
| **Random effects model** | | | **235** | **230** | | **−0.23** | **[−0.57; 0.11]** | **100.0%** |

Heterogeneity: $I^2 = 0\%$, $\tau^2 = 0$, $p = 0.49$

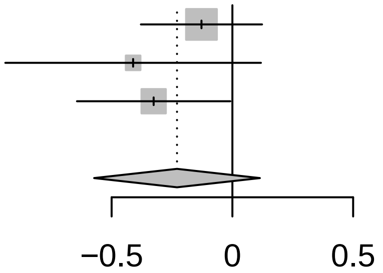
