## Supplementary material for "Animals in higher education settings: Do animal-assisted interventions improve mental and cognitive health outcomes of students? A systematic review and meta-analysis": S17_Figure

| Study | TE | seTE | N(i) Total | N(c) Total | SMD | 95%−CI |
|---|---|---|---|---|---|---|
| McDonald (2017) − systolic | −2.50 | 0.3890 | 22 | 26 | −2.50 | [−3.26; −1.74] |
| Wilson (1987) − systolic | 0.21 | 0.2100 | 46 | 46 | 0.21 | [−0.20; 0.62] |
| Crump (2015) − Study I − systolic | 0.84 | 0.4020 | 27 | 27 | 0.84 | [ 0.05; 1.63] |

Heterogeneity: $I^2 = 96\%$, $\tau^2 = 2.3304$, $p < 0.01$

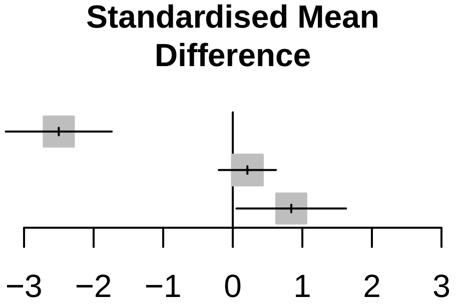
